# Nationwide population-based infection- and vaccine-induced SARS-CoV-2 seroprevalence in Germany at the end of 2021

**DOI:** 10.1101/2023.10.30.23297594

**Authors:** Elisabetta Mercuri, Lorenz Schmid, Christina Poethko-Müller, Martin Schlaud, Cânâ Kußmaul, Ana Ordonez-Cruickshank, Sebastian Haller, Ute Rexroth, Osamah Hamouda, Lars Schaade, Lothar H. Wieler, Antje Gößwald, Angelika Schaffrath Rosario, the RKI-SOEP-2 Study Group

**Affiliations:** Robert Koch Institute (RKI); Socio-Economic Panel (SOEP) at the German Institute for Economic Research (DIW Berlin); Federal Office for Migration and Refugees (BAMF); Institute for Employment Research (IAB); Digital Global Public Health at Hasso Plattner Institute (HPI); Max Planck Institute for Infection Biology (MPI)

## Abstract

**Background:** The first wave of the Corona Monitoring Nationwide (RKI-SOEP) Study drawn from the German Socio-Economic Panel proved a low pre-vaccine SARS-CoV-2 seroprevalence in the German adult population of 2.1%.

**Methods:** In this second wave of the study (RKI-SOEP-2, November 2021-March 2022), we used combined serological and self-reported data on infection and vaccination to estimate the prevalence of SARS-CoV-2-specific anti-spike and/or anti-nucleocapsid IgG antibodies (combined seroprevalence), past infection, and basic immunization in individuals aged 14+.

**Findings:** Combined seroprevalence was 90.7% (95% CI 89.7% - 91.6%) without correction for antibody waning and 94.6% (95% CI 93.6% - 95.7%) with correction. While 1 in 10 individuals had been infected (9.9%, 95% CI 9.0% - 10.9%), 9 in 10 had at least a basic immunization (90%, 95% CI 88.9%-90.9%). Population-weighted estimates differed by age, region, and socioeconomic deprivation. Infection-induced seroprevalence with correction for antibody waning was 1.55 (95% CI 1.3 - 1.8) times higher than the cumulative proportion based on national surveillance data.

**Interpretation:** At the beginning of the SARS-CoV-2-Omicron wave, the vast majority of the population had been vaccinated, infected, or both. Our results show how large-scale vaccination, but not a high infection rate, was able to fill the immunity gap, especially in older individuals (aged 65+) who are known to be at higher risk of severe COVID-19. Our data point towards a targeted demographically and regionally stratified mitigation strategy, to optimize future pandemic mitigation efforts.

## Introduction

According to national surveillance data (based on mandatory COVID-19 cases notification and vaccination monitoring), as of 30 December 2021, approximately 7,2 million people had been infected with SARS-CoV-2 in Germany and 59,2 million already had a basic immunization (based on vaccination and known infection before vaccination). However, uncertainty remains about the true exposure state of the population, especially given the unknown proportion of SARS-CoV-2 infections with mild or asymptomatic course that are not notified^1^. Studies have shown that previous symptomatic or asymptomatic SARS-CoV-2 infection alone may not be sufficient to prevent reinfection and COVID-19 disease, especially in older individuals (>65 years)^2^. Rather, partial protection against infection and strong and sustained protection against severe disease can be conferred by repeated exposures to the virus antigen through vaccinations^3^ or the combination of infection and vaccination (hybrid immunity) which offers the highest magnitude and durability of protection^4-7^. However, national surveillance data on infection and vaccination could not easily be combined in Germany, leading to uncertainty about population hybrid immunity.

The first Corona Monitoring Nationwide (RKI-SOEP) Study had shown that shortly before the start of the German immunization programme on 27.12.2020, only 2.1% of adults (18+ years) had already experienced SARS-CoV-2 infection, proving an efficient containment strategy. More than half of these cases had been detected and notified^8^. More studies have been published in Germany on SARS-CoV-2 seroprevalence. However, apart from the GUIDE study^9^, which limits its analysis to the adult population (18+ years), such studies focused on a regional or local level^10^ or were limited to specific subpopulations or settings such as blood donors^11,12^, health or educational institutions^13,14^. The second Corona Monitoring Nationwide (RKI-SOEP-2) Study, presented here, was conducted from November 2021 to February 2022 during the fourth pandemic wave in Germany that was dominated by the Delta variant, followed by the Omicron wave beginning around the turn of the year 2021-22^15^ (Figure 1). The vaccination campaign had been running for one year, and the campaign offering a booster dose had been initiated (Figure 1). In November 2021, the “2G” rule was implemented which allowed only people who had either been recovered or fully vaccinated to enter certain facilities or events. The “2G plus” rule then added the requirement of an up-to-date negative antigen test.

**Figure 1.**
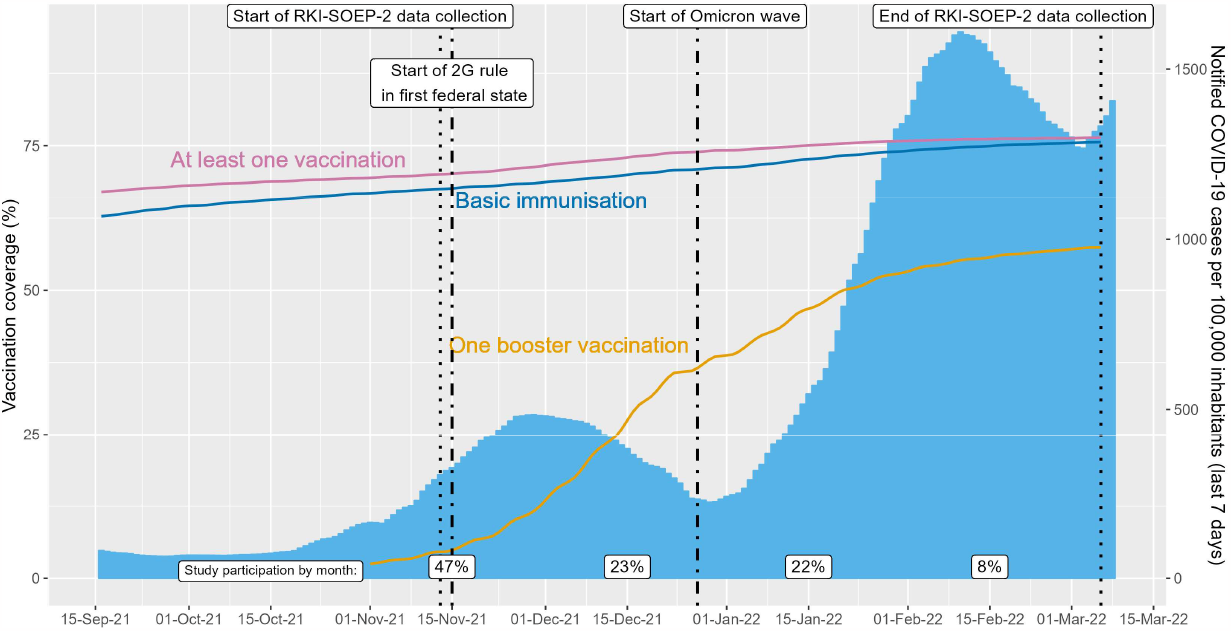
Germany, September 2021 - March 2022, all ages (including children): Percentage of individuals who received at least one vaccination dose (pink line), with at least a basic immunization (blue line), and who received one booster dose (orange line) according to the COVID-19 Digital Vaccination Coverage Monitoring (DIM); notified COVID-19 cases per 100,000 inhabitants during the last 7 days (blue bars); and percentage of participants enrolled in the Corona Monitoring nationwide (RKI-SOEP-2) study per month (white boxes at the bottom of the figure). Definition of basic immunization in the DIM differs from the definition used in this study (see Supplement 6.1). Data sources for vaccination coverage and COVID-19 cases are the Robert Koch Institute’s github repositories (https://github.com/orgs/robert-koch-institut/repositories, accessed 11 September 2023).

The aims of the study were to estimate the proportion of the population which, at the turn of the year 2021/22, (i) was seropositive for anti-SARS-CoV-2 IgG antibodies targeting both the SARS-CoV-2 nucleocapsid and the spike protein, (ii) had been infected with SARS-CoV-2, (iii) had gained a basic immunization from vaccination or the combination of infection and vaccination (hybrid immunity). Furthermore, (iv) we estimate the extent of underreporting. We report population-weighted estimates and compare results by region and sociodemographic factors. Hereby we aim to complement national COVID-19 surveillance data and deliver empirical data on both the course of the pandemic and to inform future modelling studies.

## Methods

### Study design and study population

We present cross-sectional results of the second wave of the population-based study “Corona Monitoring nationwide” (RKI-SOEP-2), a cooperative project of the Robert Koch Institute (RKI), the Socio-Economic Panel (SOEP) at the German Institute for Economic Research (DIW Berlin), the Institute for Employment Research (IAB), and the Research Center of the Federal Office for Migration and Refugees (BAMF-FZ). The study was embedded in the SOEP panel, a German nationwide dynamic cohort based on population-based random samples, and comprised individuals aged 14 years and older who participated in the 2021 SOEP wave. The study protocol has been published elsewhere^16^. Data collection began in November 2021 and ended March 6, 2022. All SOEP households were invited by mail to participate in the study. An invitation packet was sent to each respondent, which contained both the personal invitation and the study materials. To encourage participation in the study, a post-paid monetary incentive (10 euros for adults, 5 euros for adolescents) was offered to each participant as well as a written laboratory test result notification. The study was approved by the ethics committee of the Berlin Medical Association (Eth-33/20).

### Data collection and laboratory methods

Study materials included a questionnaire of twelve pages, a disposable kit for the self-sampling of capillary blood from a finger prick, illustrated instructions and a link to video material on self-sampling. The questionnaire was available in seven languages and included questions on past SARS-CoV-2 infection (previous infection detected by PCR testing, date of positive test result, severity of illness), vaccination status (number of doses, date and site of vaccination, vaccine type), attitudes toward the pandemic, health status, and health behaviours^16^. It could be answered on paper or online.

Participants were asked to send dried blood samples (DBS) by mail to the RKI laboratory. The samples were tested for SARS-CoV-2 anti-S (S1 domain of the spike protein) and anti-N (nucleocapsid protein, NCP) IgG antibodies. Anti-S antibodies were assessed with a quantitative assay, anti-N antibodies with a semi-quantitative assay (both Euroimmun AG, Lubeck, Germany). Details on measurement methods and extensive quality assessments are described in Supplement 2.1. We performed a validation study (Supplement 2.2) to check whether the manufacturer-supplied cutpoint for defining seropositivity, which was derived for serum samples, can be applied to dried blood spots. As a result, the cutpoint for anti-N seropositivity was adapted from 1.1 to 0.95, while there was no change to the cutpoint of 11 for anti-S seropositivity.

### Primary outcomes

Combined seroprevalence was defined as seropositivity for anti-N and/or anti-S antibodies. For this outcome, we present estimates both without and with correction for test characteristics (sensitivity and specificity). The estimate with correction accounts for antibody waning and thus allows estimation of the proportion of the population that has ever been exposed to SARS-CoV-2 antigens.

SARS-CoV-2 infection status was defined as ‘past infection’ when having any of self-reported infection (known PCR-confirmed infection reported in the questionnaire), seropositivity for anti-N antibodies or, if self-reported as unvaccinated, for anti-S antibodies.

Basic immunization was defined as having at least two antigen exposures, i.e. at least two self-reported vaccination doses, or a combination of at least one self-reported vaccination dose and an infection (self-reported or seropositivity for anti-N). Regardless of their chronological order, we refer to the combination of natural infection and vaccination as “hybrid immunity”. A minimum time interval was required for two subsequent events to be evaluated as separate, immunologically effective events. A more detailed description of how indicators were built is presented in Supplement 1. We do not report on three or more antigen exposures here, since the vaccination booster campaign was ongoing during field time.

Underreporting of infections in national COVID-19 surveillance data was estimated by linking the study data to surveillance data on an individual basis (by DBS sampling date, age group, sex and district). The underreporting factor was calculated as the ratio of infection-induced seroprevalence to the prevalence of notified cases (excluding deceased cases). Hereby, infection-induced seroprevalence was defined as seropositivity for anti-N antibodies, with correction for test characteristics. We use this indicator for the estimation of underreporting as it accounts for antibody waning over time since infection (similarly to self-reported known infections), but also for previously undetected infections. Details can be found in Supplement 5.

### Covariable definitions

Results are presented stratified by five age groups (14-17, 18-34, 35-49, 50-64, 65-99 years), sex, and place of residence. The latter was classified, firstly, into three categories of district-level socio-economic deprivation (low: quintile 1, medium: quintiles 2–4, high: quintile 5), and secondly, into four regions based on the federal states. Socioeconomic deprivation was measured at the level of Germany’s 400 districts using the German Index of Socioeconomic Deprivation (GISD)^17,18^ which measures relative deprivation in the domains of education, employment and income. Federal states were grouped into an eastern (Berlin, Brandenburg, Saxony, Saxony-Anhalt, Thuringia), western (North Rhine-Westphalia, Hesse, Rhineland-Palatinate, Saarland), southern (Bavaria, Baden-Württemberg) and northern region (Schleswig-Holstein, Bremen, Hamburg, Lower Saxony, Mecklenburg-Pomerania). These regions reflect the geography as well as the incidence of SARS-CoV-2 infections in the mandatory surveillance system over the course of the pandemic.

### Statistical analyses

All analyses were weighted to allow the generalization of our findings to the general population and to adjust for survey non-response. The weighting factors resulted from complex modelling of contactability and participation probabilities at both the household and the individual level. Furthermore, the weights were adjusted both at the household (household typology, size, home ownership, and federal state) and at the individual level (age, sex, and citizenship) to match the population distributions provided by the Federal Statistical Office. Details of the weighting and sampling methods have been published elsewhere^16^.

Descriptive results are presented as unweighted numbers and population-weighted percentages, for the total population as well as stratified by sex, age group, district-level socio-economic deprivation and region. 95% confidence intervals (CI) were calculated on the logit scale to ensure a value between 0 and 100%. CIs were based on robust standard errors calculated via survey procedures to account for weighting and household clustering. Logistic regression models were estimated for the primary outcomes to obtain adjusted odds ratios, also using survey procedures. The models included the four stratification variables as covariates, as well as the month of study participation (categorical variable) as control variable. p-values for the model covariates were obtained from a Wald test based on robust standard errors. Based on the logistic models, average predicted prevalences, adjusted for the covariates, were estimated with the Stata ‘margins’ command with 95% logit CIs using the command ‘coef_table’. Analyses were performed with Stata 16.1 (StataCorp, 2019, College Station, TX, USA) and SAS 9.4 (SAS Institute Inc., Cary, NC, USA).

### Correction of seroprevalence estimates for test characteristics

Both combined seroprevalence and infection-induced seroprevalence, but not infection status and basic immunization, were corrected for test characteristics using the formula^19^

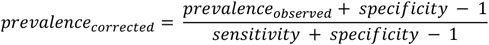

For specificity, we used the values determined for the Euroimmun assays by the Paul Ehrlich Institute^20^: 99.4% (95% CI 98.5 - 99.8) for the anti-S assay and 99.3% (95% CI 98.3 - 99.8) for the anti-N assay. For the combined seroprevalence, we used the anti-S specificity, as the proportion of participants who were anti-S seronegative but anti-N seropositive was small (0.3%). Sensitivity was estimated internally from the study population, i.e. we estimated (a) the proportion that was seropositive for combined seroprevalence (95.8%, 95% CI 95.1 - 96.4) among study participants with a self-reported vaccination or a self-reported positive SARS-CoV-2 test at least 11 days pre-study (n = 9,260); and (b) the proportion that was seropositive for anti-N antibodies (47.4%, 95% CI 41.6 - 53.3) among participants with a self-reported positive SARS-CoV-2 test at least 11 days pre-study (n = 774). For stratified analyses, we estimated sensitivity within the same strata that were used for the analysis (see Supplement 3) and performed the correction within each stratum. As a robustness check, we used additional stratification by the number of vaccinations received when estimating sensitivity, but results were not altered materially (data not shown). Confidence intervals for the corrected seroprevalence were obtained as Wald intervals taking the variability in the estimation of the stratum-specific sensitivity into account via the delta method^19^. In the logistic regression models, we corrected for test characteristics using predictive value weighting (see Supplement 4).

## Results

The study included 11,162 participants aged 14-99 years from 6,760 households. 20,774 individuals aged 14 years and older from 11,785 households had been invited to participate (response 53.7%^16^). Data were collected predominantly between November 2021 and January 2022 (Figure 1). DBS specimens yielding valid laboratory test results were available from 10,687 participants (95.7% of all participants). Questionnaire items on prior vaccination and known infection were answered by 10,985 participants (Figure 2).

**Figure 2.**
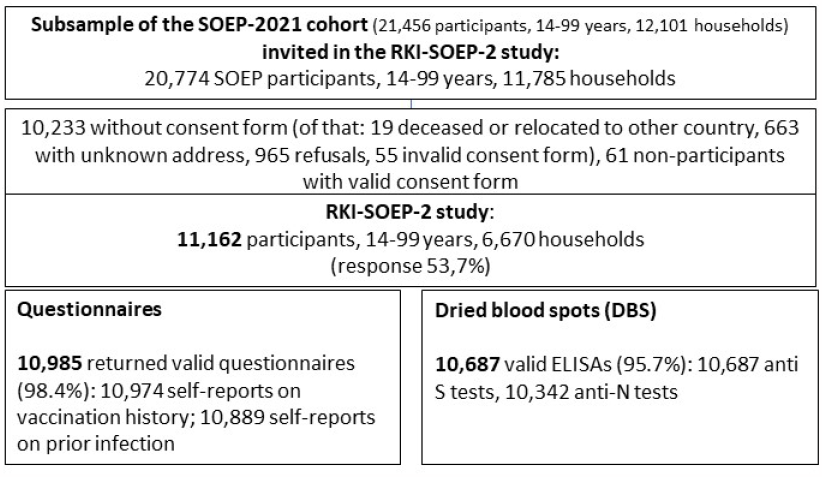
Flow-chart of the study design. Percentages are unweighted.

Tables 1, 2 and 3 present the socio-demographic characteristics of participants included in the analysis of combined seroprevalence (seropositive for anti-N and/or anti-S antibodies), past SARS-CoV-2 infection and basic immunization, respectively. The tables also show the population-weighted prevalence estimates for the primary outcomes, followed by the adjusted odds ratios (OR) and the model-adjusted prevalence estimates derived from the logistic regression models. Model-adjusted prevalence estimates differed only slightly from unadjusted estimates. Figure 3 visualizes the prevalence estimates by age group.

**Table 1.** Characteristics and combined IgG seroprevalence in community-dwelling persons (≥14 years) in Germany (10,687 RKI-SOEP-2 study participants with valid dried blood spot specimens, sampled predominantly between November 2021-January 2022).

|  | Total | N+* | S+* | Combined Seroprevalence (N+ or S+), uncorrected |  | Combined Seroprevalence (N+ or S+), corrected for specificity and stratum-specific sensitivity |  |  |
| --- | --- | --- | --- | --- | --- | --- | --- | --- |
|  | N (column %) | %** | %** | N positive | %** | Prevalence***, % (95% CI) | OR, adjusted**** (95% CI) | Model-adjusted**** prevalence, % (95% CI) |
| All (total 14-99 years) | 10687 | 6.0% | 90.4% | 9795 | 90.7% (89.7 - 91.6) | 94.6% (93.6 - 95.7) |  |  |
| All (total 18-99 years) | 10047 | 5.8% | 90.9% | 9253 | 91.2% (90.3 - 92.1) | 95.5% (94.4 - 96.6) |  |  |
| **Sex** |  |  |  |  | *p* = 0.76 |  |  |  |
| Women | 5768 (51.1) | 5.3% | 90.6% | 5300 | 90.8% (89.6 - 91.9) | 94.3% (92.8 - 95.7) | Ref. | 94.1% (92.9 - 95.1) |
| Men | 4919 (48.9) | 6.7% | 90.2% | 4495 | 90.6% (89.2 - 91.8) | 95.0% (93.3 - 96.8) | 1.30 (0.96 - 1.78) | 95.3% (94.2 - 96.3) |
| **Age group** |  |  |  |  | *p* = 0.0007 |  |  |  |
| 14-17 years | 640 (7.9) | 8.2% | 84.3% | 542 | 84.4% (79.6 - 88.2) | 84.8% (80.4 - 89.1) | 0.34 (0.21 - 0.53) | 83.9% (79.4 - 88.0) |
| 18-34 years | 1869 (22.6) | 6.2% | 91.3% | 1727 | 91.9% (89.6 - 93.7) | 94.3% (91.8 - 96.8) | Ref. | 93.7% (91.9 - 95.4) |
| 35-49 years | 2387 (20.8) | 7.9% | 89.7% | 2177 | 90.0% (87.6 - 92.0) | 93.1% (90.5 - 95.7) | 0.92 (0.60 - 1.43) | 93.2% (91.2 - 95.0) |
| 50-64 years | 3387 (25.6) | 4.5% | 92.1% | 3142 | 92.1% (90.6 - 93.4) | 96.1% (94.2 - 98.0) | 1.78 (1.12 - 2.73) | 96.3% (95.2 - 97.4) |
| 65-99 years | 2404 (23.0) | 4.7% | 90.5% | 2207 | 90.7% (88.9 - 92.3) | 98.2% (95.7 - 100.8) | 4.22 (2.23 - 6.94) | 98.4% (97.4 - 99.0) |
| **Socio-economic deprivation** |  |  |  |  | *p* = 0.0005 |  |  |  |
| Low | 2585 (25.9) | 6.3% | 92.6% | 2414 | 93.1% (91.4 - 94.5) | 96.5% (94.6 - 98.5) | Ref. | 96.5% (94.6 - 97.9) |
| Medium | 6515 (60.0) | 5.6% | 90.3% | 5978 | 90.4% (89.1 - 91.6) | 94.1% (92.6 - 95.7) | 0.59 (0.32 - 1.06) | 94.4% (93.2 - 95.4) |
| High | 1487 (14.1) | 6.9% | 86.8% | 1310 | 87.1% (83.9 - 89.7) | 93.3% (89.4 - 97.2) | 0.46 (0.21 - 0.96) | 92.9% (89.6 - 95.4) |
| **Region** |  |  |  |  | *p* < 0.0001 |  |  |  |
| Northern | 1960 (17.8) | 3.8% | 89.9% | 1812 | 89.9% (87.3 - 92.1) | 95.1% (91.8 - 98.5) | 0.58 (0.35 - 1.12) | 95.0% (93.0 - 96.9) |
| Western | 3368 (34.7) | 5.3% | 92.2% | 3145 | 92.3% (90.6 - 93.6) | 96.6% (94.6 - 98.5) | Ref. | 97.0% (95.7 - 97.9) |
| Southern | 3001 (29.5) | 6.4% | 91.4% | 2763 | 92.0% (90.3 - 93.5) | 95.3% (93.4 - 97.3) | 0.54 (0.28 - 1.02) | 94.6% (92.0 - 96.4) |
| Eastern | 2358 (18.1) | 8.8% | 86.0% | 2075 | 86.3% (83.6 - 88.7) | 89.5% (86.7 - 92.3) | 0.29 (0.18 - 0.47) | 90.6% (88.2 - 92.9) |

Numbers do not add up to total due to missing values in single variables (available-case analysis).
All analyses for 14-99 years unless otherwise specified.
\*N+, anti-N seropositive. S+, anti-S seropositive. Number of available cases for anti-N-antibodies result: 10,342; anti-S-antibodies result: 10,687.
\*\*Percentage without correction for sensitivity and specificity.
\*\*\*Prevalence for N+ and/or S+, corrected for sensitivity (including antibody waning, internally estimated, stratum-specific values, see Supplement 3) and anti-S specificity = 0.994.
\*\*\*\*Odds ratio mutually adjusted for the variables presented in the table and month of study participation (categorical variable). Bootstrap confidence intervals.

**Table 2.** Characteristics and SARS-CoV-2 infection status in community-dwelling persons (≥14 years) in Germany (11,154 RKI-SOEP-2 study participants, predominantly November 2021-January 2022).

|  | Total | self-reported<br>PCR+* | N+* | unvaccinated &<br>S+* | SARS-CoV-2 Infection |  |  |  |
| --- | --- | --- | --- | --- | --- | --- | --- | --- |
|  | N (column %) | % | % | % | N<br>positive | Prevalence, % (95%<br>CI) | OR and *p*-value,<br>adjusted** (95% CI) | Model-adjusted**<br>prevalence, % (95% CI) |
| All (total 14-99 years) | 11154 | 7.4 | 6.0 | 1.6 | 1193 | 9.9 (9.0 - 10.9) |  |  |
| All (total 18-99 years) | 10442 | 7.3 | 5.8 | 1.1 | 1100 | 9.8 (8.9 - 10.8) |  |  |
| **Sex** |  |  |  |  |  |  | *p* = 0.097 |  |
| Women | 5996 (50.9) | 7.0 | 5.3 | 1.3 | 651 | 9.1 (8.1 - 10.2) | Ref. | 9.2 (8.2 - 10.3) |
| Men | 5158 (49.1) | 7.9 | 6.7 | 1.9 | 542 | 10.8 (9.5 - 12.2) | 1.16 (0.97 - 1.38) | 10.5 (9.3 - 11.9) |
| **Age group** |  |  |  |  |  |  | *p* < 0.0001 |  |
| 14-17 years | 712 (8.4) | 8.3 | 8.2 | 6.9 | 93 | 10.9 (8.0 - 14.7) | 0.96 (0.65 - 1.42) | 10.4 (7.7 - 14.0) |
| 18-34 years | 1932 (22.3) | 8.0 | 6.2 | 1.4 | 241 | 11.3 (9.4 - 13.5) | Ref. | 10.8 (8.9 - 13.0) |
| 35-49 years | 2477 (20.6) | 11.5 | 7.9 | 2.0 | 329 | 14.4 (12.1 - 17.0) | 1.33 (1.00 - 1.78) | 13.8 (11.7 - 16.3) |
| 50-64 years | 3487 (25.4) | 6.5 | 4.6 | 1.1 | 339 | 7.9 (6.6 - 9.4) | 0.70 (0.53 - 0.92) | 7.8 (6.6 - 9.3) |
| 65-99 years | 2546 (23.3) | 4.0 | 4.7 | 0.1 | 191 | 6.5 (5.4 - 7.9) | 0.62 (0.46 - 0.84) | 7.1 (5.9 - 8.6) |
| **Socio-economic deprivation** |  |  |  |  |  |  | *p* = 0.91 |  |
| Low | 2700 (26.0) | 7.4 | 6.3 | 1.9 | 260 | 9.6 (7.8 - 11.8) | Ref. | 10.1 (8.0 - 12.8) |
| Medium | 6779 (59.8) | 7.7 | 5.6 | 1.5 | 751 | 10.0 (8.9 - 11.3) | 0.97 (0.70 - 1.34) | 9.9 (8.7 - 11.1) |
| High | 1572 (14.3) | 5.9 | 6.9 | 1.1 | 171 | 9.6 (7.5 - 12.1) | 0.91 (0.60 - 1.39) | 9.4 (7.3 - 12.0) |
| **Region** |  |  |  |  |  |  | *p* < 0.0001 |  |
| Northern | 2030 (17.7) | 4.3 | 3.8 | 0.5 | 130 | 6.3 (4.8 - 8.2) | 0.72 (0.50 - 1.02) | 6.7 (5.1 - 8.7) |
| Western | 3520 (34.7) | 7.0 | 5.3 | 0.8 | 334 | 9.0 (7.6 - 10.7) | Ref. | 9.1 (7.5 - 10.9) |
| Southern | 3146 (29.5) | 7.4 | 6.4 | 2.2 | 342 | 10.0 (8.4 - 12.0) | 1.06 (0.76 - 1.47) | 9.5 (7.7 - 11.6) |
| Eastern | 2458 (18.0) | 11.3 | 8.8 | 3.1 | 387 | 15.0 (12.8 - 17.6) | 1.79 (1.36 - 2.36) | 15.0 (12.6 - 17.8) |

Numbers do not add up to total due to missing values in single variables (available-case analysis). All percentages are population-weighted. All analyses for 14-99 years unless otherwise specified.
\*N+, anti-N seropositive. S+, anti-S seropositive. Number of available cases for self-reported PCR test: 10,889; anti-N-antibodies result: 10,342; vaccination status & anti-S-antibodies result: 10,501.
\*\*Model variables: sex, age group, district-level socio-economic deprivation, region, month of study participation. *p*-value for a Wald test of each variable within a survey logistic regression model.

**Table 3.** Characteristics and basic immunization status in community-dwelling persons (≥14 years) in Germany (10,932 RKI-SOEP-2 study participants, predominantly November 2021-January 2022).

|  | Total | at least two doses of vaccine* | hybrid immunity* | Basic Immunization |  |  |  |
| --- | --- | --- | --- | --- | --- | --- | --- |
|  | N (column %) | % (95% CI) | % (95% CI) | N positive | Prevalence, % (95% CI) | OR and *p*-value, adjusted** (95% CI) | Model-adjusted** prevalence, % (95% CI) |
| All (total 14-99 years) | 10932 | 88.1 (87.0 - 89.1) | 7.5 (6.7 - 8.3) | 9975 | 90.0 (88.9 - 90.9) |  |  |
| All (total 18-99 years) | 10250 | 89.1 (88.0 - 90.1) | 7.8 (7.0 - 8.6) | 9439 | 91.0 (90.0 - 92.0) |  |  |
| **Sex** |  |  |  |  |  | *p* = 0.51 |  |
| Women | 5897 (51.1) | 89.2 (87.9 - 90.3) | 7.2 (6.3 - 8.2) | 4598 | 90.6 (89.3 - 91.7) | Ref. | 90.2 (89.0 - 91.4) |
| Men | 5035 (48.9) | 87.0 (85.4 - 88.4) | 7.8 (6.7 - 9.0) | 5377 | 89.3 (87.8 - 90.7) | 0.94 (0.77 - 1.14) | 89.7 (88.3 - 91.0) |
| **Age group** |  |  |  |  |  | *p* < 0.0001 |  |
| 14-17 years | 682 (8.2) | 77.1 (71.9 - 81.6) | 4.3 (2.9 - 6.5) | 536 | 78.1 (72.9 - 82.5) | 0.51 (0.36 - 0.72) | 77.3 (72.2 - 81.7) |
| 18-34 years | 1880 (22.1) | 84.6 (81.8 - 87.0) | 8.1 (6.6 - 9.8) | 1671 | 87.5 (84.8 - 89.7) | Ref. | 86.8 (84.1 - 89.1) |
| 35-49 years | 2412 (20.6) | 84.2 (81.5 - 86.6) | 11.4 (9.3 - 13.9) | 2136 | 87.7 (85.2 - 89.8) | 1.11 (0.82 - 1.50) | 88.0 (85.5 - 90.0) |
| 50-64 years | 3442 (25.6) | 90.6 (88.9 - 92.0) | 6.5 (5.3 - 7.8) | 3188 | 91.9 (90.3 - 93.3) | 1.78 (1.33 - 2.37) | 92.0 (90.5 - 93.3) |
| 65-99 years | 2516 (23.5) | 95.9 (94.4 - 96.9) | 5.7 (4.7 - 7.0) | 2444 | 96.4 (95.0 - 97.4) | 4.23 (2.78 - 6.44) | 96.5 (95.1 - 97.5) |
| **Socio-economic deprivation** |  |  |  |  |  | *p* = 0.11 |  |
| Low | 2654 (26.0) | 90.6 (88.6 - 92.3) | 7.2 (5.7 - 9.2) | 2477 | 92.1 (90.1 - 93.7) | Ref. | 92.1 (89.9 - 93.8) |
| Medium | 6645 (59.9) | 87.4 (85.9 - 88.8) | 7.5 (6.6 - 8.5) | 6035 | 89.4 (88.0 - 90.7) | 0.72 (0.52 - 1.00) | 89.4 (88.0 - 90.7) |
| High | 1535 (14.1) | 86.5 (83.2 - 89.2) | 7.8 (6.0 - 10.1) | 1371 | 88.5 (85.4 - 91.1) | 0.64 (0.40 - 1.01) | 88.3 (84.8 - 91.1) |
| **Region** |  |  |  |  |  | *p* < 0.0001 |  |
| Northern | 1987 (17.8) | 89.6 (86.8 - 91.8) | 5.0 (3.8 - 6.5) | 1911 | 90.5 (87.9 - 92.7) | 0.75 (0.52 - 1.07) | 90.6 (88.0 - 92.7) |
| Western | 3445 (34.7) | 90.4 (88.5 - 91.9) | 7.8 (6.5 - 9.4) | 3226 | 92.7 (91.1 - 94.0) | Ref. | 92.7 (91.1 - 94.1) |
| Southern | 3090 (29.6) | 89.0 (86.9 - 90.8) | 6.7 (5.3 - 8.3) | 2894 | 90.1 (88.0 - 91.8) | 0.64 (0.45 - 0.91) | 89.2 (86.6 - 91.4) |
| Eastern | 2410 (18.0) | 80.8 (77.6 - 83.6) | 10.7 (8.9 - 12.9) | 2187 | 84.0 (81.0 - 86.6) | 0.44 (0.32 - 0.60) | 85.4 (82.5 - 87.8) |

Numbers do not add up to total due to missing values in single variables (available-case analysis). All percentages are population-weighted. All analyses for 14-99 years unless otherwise specified.
\*Number of available cases for at least two doses of vaccine: 10,932; hybrid immunity: 10,925.
\*\*Model variables: sex, age group, district-level socio-economic deprivation, region, month of study participation. *p*-value for a Wald test of each variable within a survey logistic regression model.

**Figure 3.**
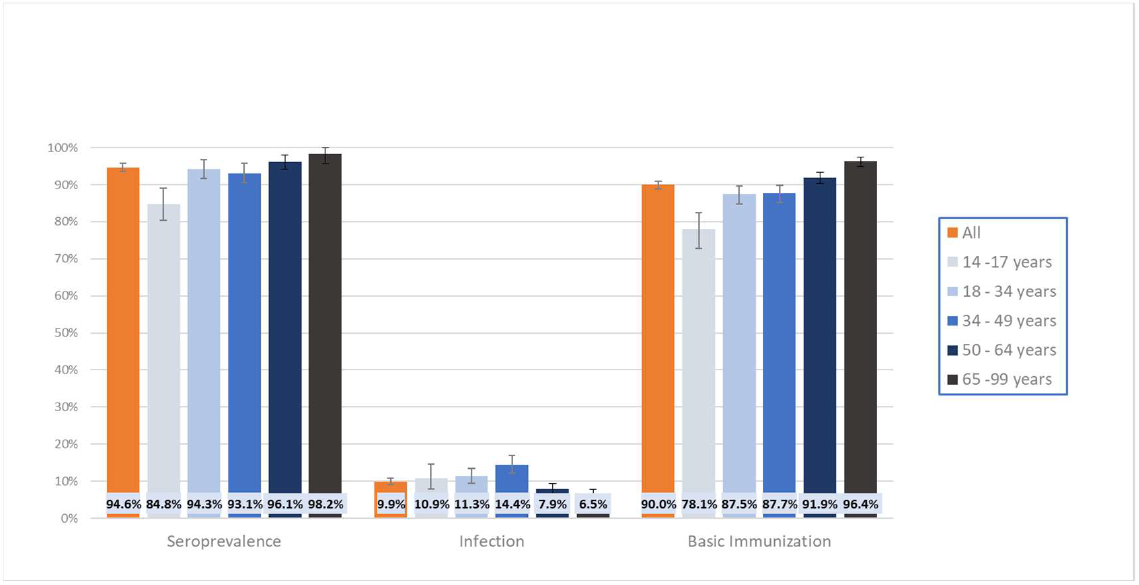
Combined seroprevalence, past SARS-CoV-2 infection and basic immunization stratified by age. All percentages are population-weighted.

Combined seroprevalence from infection, vaccination or both was 90.7% (95% CI 89.7% - 91.6%) without correction for test characteristics in the population aged 14-99 years (Table 1). Correcting the estimate for specificity and internally estimated sensitivity increased the combined seroprevalence to 94.6% (95% CI 93.6% - 95.7%). Combined seroprevalence was lowest in the youngest age group (14-17 years: 84.8%, 95% CI 80.4% - 89.1%, with correction) and in the eastern German region (89.5%, 95% CI 86.7% - 92.3%, with correction). Higher socio-economic deprivation was also associated with lower seroprevalence (93.3%, 95% CI 89.4% - 97.2%, with correction).

By combining antibody test results with information derived from the questionnaires we then estimated the prevalence of SARS-CoV-2 infection (Table 2) and basic immunization (Table 3). By the turn of the year 2021-22, 9.9% (95% CI 9.0% - 10.9%) of the population in Germany were estimated to have been infected with SARS-CoV-2 (Table 2). Infection prevalence increased with age, up until the age group 35-49 years (14.4%, 95% CI 12.1% - 17%), but was only half as high in the higher age groups (7.9%, 95% CI 6.6% - 9.4% for the age group 50-64 years and even lower at 6.5% for 65-99 years, 95% CI 5.4% - 7.9%). When comparing by geographic area, the highest infection rate was detected in the eastern region (15%, 95% CI 12.8% - 17.6%) and the lowest in the northern region (6.3%, 95% CI 4.8% - 8.2%).

The percentage of the population with a basic immunization (at least two exposures to the virus antigen) was estimated as 90.0% (95% CI 88.9% - 90.9%). The vast majority of these developed a basic immunization from vaccination, 7.5% (95% CI 6.7% - 8.3%) had a hybrid immunity (Table 3). Basic immunization increased with age and decreased somewhat with higher socioeconomic deprivation. Compared to the other regions, in the eastern region only 84.0% (95% CI 81.0% - 86.6%) of participants had a basic immunization. Notably, basic immunization was highest (96.4%, 95% CI 95.0% - 97.4%) in the age group with the lowest infection rate (65-99 years), and it was lowest in the region with the highest infection rate (eastern region). While infection and hybrid immunity (10.7%, 95% CI 8.9% - 12.9%) were highest in the eastern region, basic immunization and the proportion who had received at least two doses of vaccine (80.8%, 95% CI 77.6% - 83.6%) was lowest compared to the other regions.

In multivariable logistic regression models, combined seroprevalence was associated with age group, socioeconomic deprivation and region (see Supplement 7 for stratification by federal state). Infection status and basic immunization were associated with age group and region. There was no statistically significant association with sex, although men seemed to be at a somewhat higher infection risk than women (OR 1.16, 95% CI 0.97 - 1.38). Compared to adults aged 18-34 years, adolescents (14-17 years) were both significantly less likely to be seropositive for anti-N and/or anti-S antibodies (OR 0.34, 95% CI 0.21 - 0.53) and to have a basic immunization (OR 0.51, 95% CI 0.36 - 0.72). The odds of being seropositive were also significantly reduced for individuals from areas of high socio-economic deprivation (OR 0.46, 95% CI 0.21 - 0.96 for high vs low) and for participants from the eastern region (OR 0.29, 95% CI 0.18 - 0.47) compared to the western region. The oldest age groups where both significantly more likely to have a basic immunization (OR 1.78, 95% CI 1.33 - 2.37 and OR 4.23, 95% CI 2.78-6.44 for 50-64 and 65-99 years, respectively) and significantly less likely to have had an infection (OR 0.70, 95% CI 0.53 - 0.92 and OR 0.62, 95% CI 0.46 - 0.84, respectively). Participants from the eastern and southern regions were significantly less likely to have a basic immunization (OR 0.44, 95% CI 0.32 - 0.60 and OR 0.64, 95% CI 0.45 - 0.91, respectively) than participants from the western region. Individuals from the east were also more likely to have had an infection (OR 1.79, 95% CI 1.36 - 2.36).

Table 4 presents the results regarding underreporting and infection-induced seroprevalence. Since the sensitivity for detecting anti-N antibodies is relatively low, the uncorrected infection-induced seroprevalence (Table 1) was about doubled when sensitivity was corrected for (Table 4). The correction yielded an estimate for the proportion of the population with past infection of 11.3% (95% CI 9.1% - 13.5%), which was somewhat higher than the estimate of 9.9% based on infection status, i.e. uncorrected antibody analysis combined with self-reported known infections (Table 2). The observed patterns in the stratified analyses, however, were similar. The only exception is the ranking of the age groups 18-34 and 35-49 years: When sensitivity is controlled for (Table 4), the infection rate is highest in 18-34-year-olds, albeit a wide CI (16.2%, 95% CI 9.1% - 23.2%), at a similar level to the age group 35-49 years. When sensitivity is not controlled for, however (Table 2), the infection rate in 18-34-year-olds is lower (11.3%, 95% CI 9.4% - 13.5%) than in the age group 35-49 years and is closer to the adolescent age group.

**Table 4.** Infection-induced seroprevalence and underreporting factor in community-dwelling persons (>14 years) in Germany (10,342 RKI-SOEP-2 study participants, predominantly November 2021-January 2022).

|  | Total | Infection-induced seroprevalence (N+)*, corrected for specificity and stratum-specific sensitivity |  |  | Cumulative proportion of non-fatal cases in notification data** | Underreporting factor*** |  |  |
| --- | --- | --- | --- | --- | --- | --- | --- | --- |
|  | N (column %) | N positive | % | (95% CI) |  | Ratio | (95% CI) | p-value |
| All (total 14-99 years) | 10342 | 621 | 11.3% | (9.1 - 13.5) | 7.3% | 1.55 | (1.3 - 1.8) |  |
| All (total 18-99 years) | 9729 | 568 | 11.4% | (9.1 - 13.6) | 7.0% | 1.62 | (1.3 - 1.9) |  |
| **Sex** |  |  |  |  |  |  |  |  |
| Women | 5603 (51.2) | 331 | 10.2% | (7.4 - 13.0) | 7.3% | 1.39 | (1.0 - 1.8) | Ref. |
| Men | 4739 (48.8) | 290 | 12.4% | (9.0 - 15.9) | 7.2% | 1.72 | (1.2 - 2.2) | 0.28 |
| **Age group** |  |  |  |  |  |  |  |  |
| 14-17 years | 613 (7.9) | 53 | 11.6% | (5.5 - 17.7) | 10.3% | 1.12 | (0.5 - 1.7) | 0.27 |
| 18-34 years | 1814 (22.7) | 113 | 16.2% | (9.1 - 23.2) | 9.8% | 1.65 | (0.9 - 2.4) | Ref. |
| 35-49 years | 2350 (21.3) | 163 | 15.6% | (10.0 - 21.1) | 8.8% | 1.77 | (1.1 - 2.4) | 0.81 |
| 50-64 years | 3293 (25.8) | 177 | 8.0% | (4.9 - 11.1) | 6.1% | 1.30 | (0.8 - 1.8) | 0.42 |
| 65-99 years | 2272 (22.4) | 115 | 7.3% | (4.3 - 10.4) | 3.7% | 2.00 | (1.2 - 2.8) | 0.54 |
| **Socio-economic deprivation** |  |  |  |  |  |  |  |  |
| Low | 2502 (26.0) | 140 | 11.0% | (6.5 - 15.5) | 7.4% | 1.49 | (0.9 - 2.1) | Ref. |
| Medium | 6306 (60.0) | 381 | 11.2% | (8.2 - 14.2) | 7.3% | 1.54 | (1.1 - 2.0) | 0.89 |
| High | 1438 (14.1) | 95 | 10.6% | (6.2 - 15.0) | 7.2% | 1.47 | (0.9 - 2.1) | 0.96 |
| **Region** |  |  |  |  |  |  |  |  |
| Northern | 1910 (17.8) | 77 | 7.1% | (3.1 - 11.2) | 4.6% | 1.55 | (0.7 - 2.4) | 0.95 |
| Western | 3252 (34.5) | 165 | 10.1% | (6.3 - 14.0) | 6.7% | 1.52 | (0.9 - 2.1) | Ref. |
| Southern | 2908 (29.7) | 177 | 11.9% | (7.5 - 16.3) | 8.3% | 1.43 | (0.9 - 2.0) | 0.83 |
| Eastern | 2272 (18.0) | 202 | 16.7% | (11.2 - 22.2) | 9.5% | 1.76 | (1.2 - 2.3) | 0.56 |

Numbers do not add up to total due to missing values in single variables (available-case analysis). All percentages are population-weighted. All analyses for 14-99 years unless otherwise specified.
\*N+, anti-N seropositive. Prevalence for N+, corrected for sensitivity (including antibody waning, internally estimated, stratum-specific values, see Supplement 3) and specificity = 0.993.
\*\*Cumulative proportion of notified cases, adjusted for sampling density (i.e. each participant contributes according to the sex-, age group- and district-specific cumulative proportion of notified cases with symptom onset corresponding to his/her DBS testing date minus 14 days).
\*\*\*Underreporting factor: Ratio of infection-induced seroprevalence in the study to cumulative proportion of notified cases.

Comparing the corrected anti-N seroprevalence to notified case numbers, we found an underreporting ratio of 1.55 (95% CI 1.3 - 1.8), referring to the first two years of the pandemic. There was some suggestive evidence of higher underreporting in men compared to women and in the eastern region, while it seemed lower in 14- to 17-year-olds, but statistical uncertainty was high and no result reached statistical significance (Table 4).

## Discussion

Our study shows that by the turn of the year 2021-22, when the Omicron wave was beginning, around 19 out of 20 persons aged 14 years and older were seropositive for anti-SARS-CoV-2 antibodies in Germany. The remaining 5% of the population was still immunologically naïve. Based on self-reports and serologic evidence we estimated that around 1 in 10 individuals had been infected with SARS-CoV-2, and 9 in 10 had experienced at least two exposures to the virus antigen primarily from vaccination and to a much lesser extent (7.5%) from the combination of vaccination and infection (hybrid immunity). The underreporting factor for the first two years of the pandemic in Germany was estimated to be as low as 1.55.

The added value of our study derives from multiple aspects: (i) it is based on a representative nationwide sample, so data are generalizable to the general population, (ii) information collected from participants is very detailed, (iii) analyses of seroprevalence were corrected for antibody waning, (iv) we provide an estimate of underreporting, (v) the study includes adolescents, who showed prevalence patterns different from adults, and (vi) it was possible to assess the prevalence of hybrid immunity, which we could define at the individual level thanks to the combined analysis of serological and questionnaire data.

There are two other studies reporting nationwide estimates of vaccination- and infection-induced seroprevalence in Germany, the SeBluCo study^11^ in adult blood donors and the GUIDE^9^ study in the general adult population. While our study focuses on the Delta-to-Omicron transition period, SeBluCo depicts different time points since the start of the pandemic in 2020, including the beginning of the Delta-dominated fourth pandemic wave and part of the Omicron-driven fifth wave^15^. Between September 2021 and April 2022, SeBluCo showed an increase in combined seroprevalence from 89.4% to 100%, while our study showed a seroprevalence of 95.5% for adults in between those two time points, which shows a consistent development over time. In summer 2022, the GUIDE study found a seroprevalence of 95.7% for anti-S antibodies and 44.4% for anti-N antibodies (both not corrected for test characteristics), which indicates a further consistent rise of anti-S antibody prevalence (compared to uncorrected 90.9% anti-S seroprevalence among adults in our study) and a strong increase in infections during the Omicron wave. Our results can also be compared to a modelling study^21^ of notified infections and vaccinations, which yielded a pre-Omicron proportion of immunologically naïve of 10.8% (18-59 years) and 4.8% (60+ years). These estimates are higher than those obtained from our data (5.5% and 2.2% for 18-59 years and 60+ years). The discrepancy may be due to inaccuracies in notified vaccinations and to idealized assumptions in the modelling study, but it also indicates a possible overrepresentation of vaccinated individuals in our study due to selection bias, as discussed in Supplement 6.1.

A strength of seroprevalence studies is the possibility to estimate the extent of underreporting in mandatory COVID-19 case notification. We found an underreporting factor of 1.55 (95% CI 1.3 - 1.8) which refers to the first two years of the pandemic combined. This factor is lower than the one we observed after the first pandemic year, which was 1.82 (95% CI 1.3 - 2.5)^8^. This decrease in underreporting over time is consistent with other studies from Germany^1,11^ and can be explained by intensified diagnostics and the improved access to SARS-CoV-2 tests over time. As in the first wave of the study, we found suggestive evidence of higher underreporting in areas with higher infection rates. The SeBluCo study^11^ also found higher underreporting in the eastern region, but confined to the beginning of 2021 and accompanied by higher underreporting also in the southern region. In the first wave of our study, there was suggestive evidence of higher underreporting in areas with high socio-economic deprivation. This finding was not replicated in the present analysis, indicating that regional inequalities in access to testing may have been reduced over the course of the pandemic.

Consistent with previous findings showing disparities in both vaccination coverage and infection rates across different demographic groups^25-27^, we found that combined seroprevalence differed by age, geographic area and district-level socio-economic deprivation, while infection and basic immunization differed by age and geographic area. The still relatively low infection rate recorded in the overall population is most probably due to the fact that the majority of our data were collected just before the more transmissible Omicron variant became dominant over the Delta variant^28^. The finding of higher infection rates, but lower basic immunization rates, among younger age groups reflects higher incident infections reported among younger than older adults during the Delta surge^29^ and higher vaccination coverage among older adults. Higher infection rates were detected in the eastern region, which was also characterized by the lowest prevalence of basic immunization. These empirical findings on regional differences, which were also obtained in the GUIDE study^9^, support national surveillance data. Importantly, our data highlight how the higher prevalence of infection and of hybrid immunity detected in the eastern region is not sufficient to counterbalance the low prevalence of two vaccine doses when it comes to achieving a basic immunization. This corresponds to findings of a modelling study demonstrating how a minority of the German population, the unvaccinated, were the primary driver of the rise in new infections in the fall of 2021^30^.

One of the major strengths of our study is the use of a nationwide, population-based sample covering all the 400 districts in Germany, drawn from a long-running dynamic cohort that allows for sophisticated weighting and thus higher generalizability to the general population. Although some selection bias may be present, in general our study population shows good comparability with national surveillance data (see Supplement 6). The second strength of our study is the high quality of the data collected. Participants were asked to provide detailed information on their infection and vaccination status which enhances the validity of the self-reports and allowed us to verify the time since last exposure as well as the time intervals between immunologically effective events. Furthermore, information collected in this population-based sample can be used for future analyses exploring different impacts of the pandemic (e.g. Long COVID). By combining information on self-reported infections and vaccinations with antibody test results, and correcting for sensitivity as estimated from the study population, we minimized the effects of antibody waning, which is particularly problematic for antibodies against the nucleocapsid of SARS-CoV-2. Still, the analysis depends on the assumption that test sensitivity is equal for known and unknown infections (within strata). Sensitivity can be expected to be lower in asymptomatic and mild infections^22,23^, so there might be some underestimation of the anti-N seroprevalence and the underreporting factor. Finally, our study provides a nationwide estimate on the proportion of the population with hybrid immunity, which reached 7.5% before the Omicron wave. Hybrid immunity resulting from vaccination in addition to SARS-CoV-2 infection before or after vaccination provides more robust immune response and increased protection from infection and severe disease compared with either vaccination or infection alone^4-7,24^.

Our findings must be interpreted in light of the various potential sources of bias including the underrepresentation of specific settings with increased infection rates, such as long-term care facilities, which may have undermined the generalizability of our findings. The selective participation of more health-conscious individuals may have led to some overestimation of the proportion of vaccinated individuals (see Supplement 6.1). At the same time, the proportion immunologically naïve, the proportion of individuals with past infection and the underreporting factor will be somewhat underestimated. Since no characteristics directly related to the pandemic such as attitudes toward anti-pandemic measures could be included in the weighting process, this selective participation could not be entirely corrected for. Some participation bias regarding known infections was found in the younger age groups, where individuals with known infections were underrepresented in the age groups 14-17 and 18-34 years, and overrepresented in the age group 35-49 years (see Supplement 6.2). Observed patterns in our study comparing infection rates between the age groups 18-34 years and 35-49 years are therefore uncertain.

## Conclusion

Our cumulative data show that by the turn of the year 2021-22, the vast majority of the German population aged 14 years and older had been vaccinated, infected or both. Most had at least a basic immunization which was primarily due to vaccination and to a much lower extent to hybrid immunity, reflecting the successful containment strategy. Our results show how large-scale vaccination, but not a high infection rate, was able to fill the immunity gap especially in older individuals (over 65 years) who are known to be at higher risk of severe COVID-19. However, a non-negligible proportion of adolescents (around 15%), of young adults (around 6%) and of middle-aged adults (around 7%) had not been exposed to SARS-CoV-2 antigens at the start of the Omicron wave. It will be of utmost importance to address demographic and regional disparities when establishing prevention strategies in the future, including measures to enhance vaccination uptake.

## Data Availability

The data cannot be made publicly available because informed consent from participants did not cover public deposition of data. However, the dataset underlying the analysis in this article is archived in the SOEP Research Data Centre (https://www.diw.de/en/diw_01.c.601584.en/data_access.html) in Berlin and can be accessed on site upon reasonable request.

## Author contributions

AG, LaS and LHW initiated the study and acquired funding. AG administrated the project. EM coordinated the conceptualization of outcome variable definitions and manuscript, with contributions from AG, CPM, ASR, LoS and AOC. Laboratory analyses were conceptualised and supervised by MS. SH, LaS and LHW contributed to study supervision. Study data management, data validation and statistical analysis were conceptualised and performed by ASR and LoS. UR and OH conceptualised and supervised the collection of national surveillance data. Visualisation was done by LoS. EM, SH, CPM, SH, CK, AOC contributed to the literature research. The original draft was written by EM, with contributions from ASR, LoS and CK. Data interpretation, review and editing of the manuscript was done by EM, LoS, CPM, MS, CK, AOC, SH, UR, OH, LaS, LHW, AG, ASR. All authors have read and approved the final version of the manuscript.

## Acknowledgement

We acknowledge the dedicated work of the many colleagues who contributed to this study. The RKI-SOEP-2 Study Group comprises the following colleagues (all RKI, if not otherwise specified): M. Grabka (DIW), S. Zinn (DIW), H. W. Steinhauer (DIW), M. Siegert (BAMF), K. Tanis (BAMF), W. Niehues (BAMF), N. Rother (BAMF), L. Goßner (IAB), H. Brücker (IAB), P. Trübswetter (IAB), A. Schaffrath Rosario, L. Schmid, E. Mercuri, M. Schlaud, U. Kubisch, C. Kußmaul, S. Stahlberg, A. Kneuer, A. Sandoni, J. Lücke, M. Stimpel, R. Erel, M. Joch, S. Öztürk, F. Hagedorn, B. Hecke, R. Klang, W. Zhuang, J. Hofmann, A. Männel, S. Jordan, C. Poethko-Müller, J. Hoebel, S. Bartig, H. Butschalowsky, A. Gößwald, J. Wernitz, S. Torregroza (infas), D. Hess (infas). Sequence analyses by Stefan Junker and Felicitas Vogelgesang supported our grouping of the federal states for analysis. Finally, we sincerely thank all study participants for their willingness to participate.

