## Supplemental Material for "Nationwide population-based infection- and vaccine-induced SARS-CoV-2 seroprevalence in Germany at the end of 2021"

#### 1. Definition of outcome variables

Supplemental Table S1 illustrates which indicators were considered for defining each outcome variable. Combined seroprevalence and infection-induced seroprevalence were defined solely based on antibody test results. SARS-CoV-2 infection status and basic immunization were defined based on the combination of information provided in the questionnaire and antibody test results.

Only participants with at least one available information on anti-S or anti-N IgG antibody test results were included in the analysis of combined seroprevalence. Any of self-reported positive PCR test, anti-N IgG antibody test result, or anti-S IgG antibody test result had to be available to be included in the analysis of SARS-CoV-2 infection status. The latter, the anti-S antibody test result, was only included for participants who reported never having been vaccinated. Available information on at least the vaccination status was required to be included in the analysis of basic immunization. Sensitivity analyses requiring complete data yielded similar results.

| | anti-Sars-CoV-2 tot IgG antibodies | | self-reported | self-reported | time-interval |
|---|---|---|---|---|---|
| | S | N | vaccination | infection | between events |
| **Combined seroprevalence** | +/- | +/- | | | N |
| **Infection** | +/- | +/- | Y/N | Y/N | N |
| **Basic immunization** | | +/- | Y | Y/N | Y |

**Supplemental Table S1:** Indicators included in the definition of each outcome variable. Combined seroprevalence was defined as anti-N and/or anti-S seropositivity. Infection was defined based on self-reported positive PCR test or anti-N seropositivity. Participants with anti-S+ and anti-N- were also included in this category when they reported not having been vaccinated or infected. This assumed that in the absence of reported vaccination, anti-S seropositivity was an indicator of previous infection despite having no anti-N seropositivity. Basic immunization was defined as having a self-report of at least one vaccine dose and at least one additional exposure from vaccination (self-report) or infection (self-report or anti-N+). The time interval between immunologic events was evaluated to define basic immunization.

Participants who reported a previous infection detected by PCR testing in the questionnaire were defined as participants with known infection. Participants with unknown infection were defined as those with no self-report of a positive PCR test, but positive for anti-N antibodies (independently of vaccination status) as well as those with no self-report of a positive PCR test and who reported never having been vaccinated, but were positive for anti-S antibodies (see Supplementary Figure S1).

To define basic immunization, we counted the number (one, two, three, or four) of exposures to virus antigens as shown in Supplementary Figure S1. We considered individuals as having developed a basic immunization if they had at least two exposures (from two or three doses of vaccine alone or from the combination of vaccination and infection). Based on the recommendations provided by the German National Standing Committee on Vaccination (STIKO)[1,2] at that time, we evaluated a vaccination (self-reported) and a subsequent infection (self-reported) as separate immunologically effective events only if a minimum time distance occurred between the two (Supplementary Figure S1). This distance was at least four weeks if the exposure from infection occurred after the first dose of vaccine, and at least three months if the infection occurred as third exposure after a second dose of vaccine. We assumed that the minimum time interval required between two subsequent doses of vaccine as well as between vaccination and prior infection was met as recommended by national immunization guidelines at that time. A positive anti-N antibody test was considered a sign of an immunologically effective infection, even if the time requirements were not met. Anti-S antibody tests in the unvaccinated were not considered here, since a positive anti-S test would only indicate past infection and thus not fulfil the requirement of at least two antigen exposures. Only vaccines authorized by the European Medicines Agency (EMA) at the time of the study were considered (Comirnaty, Spikevax, Vaxzevria, JCOVDEN). Individuals vaccinated with JCOVDEN (previously COVID-19 Vaccine Janssen) were categorized as having a basic immunization only if they had at least a second exposure from vaccination with an mRNA-based vaccine or infection. Of note, the definition of basic immunization in the study differs somewhat from the definition in the national vaccination monitoring, since there is more information available in the study (see Supplement 6.1).

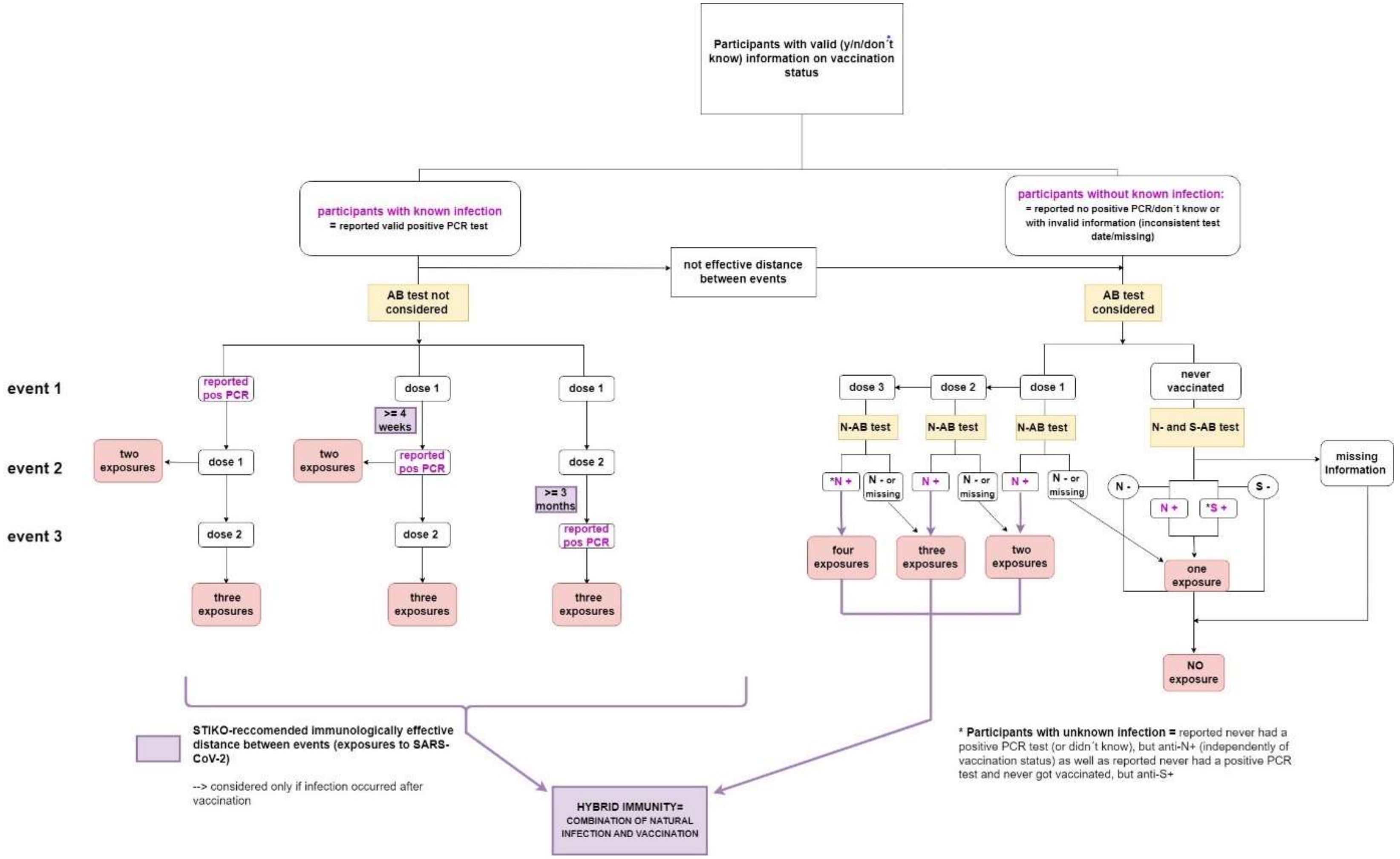

**Supplemental Figure S1:** Flowchart of the definition of basic immunization. To define basic immunization, we counted the number (one, two, three, or four) of exposures to the virus antigens, both from vaccination alone as well as from vaccination and infection (hybrid immunity). Individuals were categorized as having a basic immunization if they had at least two exposures. See main text above for detailed explanations. Distance between events was evaluated only if the infection occurred after the vaccination. We assumed indeed that the minimum time interval required between prior infection and vaccination (as well as between vaccine doses) was met as recommended by national immunization guidelines at that time. The distance required for two subsequent immunologic events to be effective, was of at least four weeks if the infection occurred after the first dose of vaccine, and at least three months if the infection occurred after the second dose of vaccine.

### 2. Detailed information on laboratory measurements

#### 2.1 Laboratory methods and quality assessment

Participants were asked to send dried blood samples (DBS) by mail to the Epidemiological Central Laboratory of the RKI. Standardized punches of DBS (DBS Puncher, PerkinElmer, Waltham MA, USA) were then extracted according to the manufacturer's protocol (Euroimmun AG, Lübeck, Germany) and tested for SARS-CoV-2 anti-S1 (S1 domain of the spike protein) and anti-N (nucleocapsid protein, NCP) IgG antibodies. The Euroimmun ELISAs (enzyme-linked immunosorbent assays) anti-SARS-CoV-2-QuantiVac (QuantiVac) and anti-SARS-CoV-2-NCP (NCP) were used, respectively, for the quantitative detection of anti-S and for the semi-quantitative detection of anti-N antibodies. For defining seropositivity for anti-N antibodies, the ratio cutpoint provided by the manufacturer for serum samples was adapted from 1.1 to 0.94 to account for the use of dried blood spots (see Supplement 2.2.). All analyses were performed on a EUROLabWorkstation ELISA (Euroimmun AG, Lübeck, Germany), testing three quality control specimens (two provided by the manufacturer and one DBS control) on each 96-well plate. The Epidemiology Laboratory at the RKI is accredited according to DIN EN ISO 17025 and DIN EN ISO 15189 (Deutsche Akkreditierungsstelle, Frankfurt/Main, Germany). Internal quality assessments were conducted for both assays. For the quality control specimens used, intra-assay coefficients of variability (CV) ranged from 4.0% to 8.2% for the QuantiVac and 6.3% to 6.4% for the NCP. Inter-Assay CVs ranged from 2.9% to 4.8% for the QuantiVac and 4.9% to 9.7% for the NCP. Relative root mean squared error (RRMSE) was used as a measure of accuracy and ranged from 4.4% to 18.5% for the QuantiVac and from 8.0% to 10.7% for the NCP. The laboratory participated in external quality assessments (EQAs) on the detection of SARS-CoV-2 IgG antibodies, offered by INSTAND interlaboratory comparison program (INSTAND, Düsseldorf, Germany) and passed all round robin tests on SARS-CoV-2 IgG antibodies.

#### 2.2 Application of the Euroimmun Anti-SARS-CoV-2-NCP (IgG) and Anti-SARS-CoV-2-QuantiVac-ELISA (IgG) antibody tests to dried blood spots

A previous validation study during the first wave of the RKI-SOEP-study revealed an adapted cutpoint of 0.94 for classifying semiquantitative values of the Euroimmun Anti-SARS-CoV-2-S1-IgG ELISA antibody test in dried blood spot (DBS) samples [3]. For the second wave of the study, different test assays were used. S-antibodies were analysed quantitatively (as opposed to the semiquantitative test in the first wave) using the Anti-SARS-CoV-2-QuantiVac-ELISA (IgG) by Euroimmun. N-antibodies, which could not yet be tested for in the first wave, were analysed semiquantitatively with the Anti-SARS-CoV-2-NCP (IgG) (also by Euroimmun). These tests are commonly used in the analysis of serum samples and are CE certified for use on DBS. However, we conducted an additional validation study comparing serum with DBS results in May and June 2022 to potentially optimize the cutpoint for DBS. A total of 244 employees of the RKI volunteered to take part in the study after an institute-wide call was published via email. Blood specimens were taken after participants' informed consent. The samples for all 244 participants could be evaluated. Results were reported back to participants anonymously and no additional information, e.g. on their age, sex, vaccination or infection history, was collected.

##### Study execution and laboratory methods

From each participant, the study team collected both a venous blood sample, which was processed into serum, and a capillary blood sample, which was processed into DBS. Both samples were then tested for IgG antibodies using the Anti-SARS-CoV-2-QuantiVac-ELISA (IgG) and Anti-SARS-CoV-2-NCP (IgG) (both by Euroimmun AG, Lübeck, Germany). The results of the S-antibody test were quantitative, expressed in binding antibody units (BAU/mL) and classified for serum samples according to the manufacturer's specifications (positive: ≥ 35.2 BAU/mL, indeterminate ≥ 25.6 to < 35.2 BAU/mL, negative: < 25.6 BAU/mL). The results of the N-antibody test were semiquantitative ratio values which were classified for serum samples using the manufacturer-supplied cutpoints (positive: ratio ≥ 1.1; indeterminate: 0.8 ≤ ratio< 1.1, negative: ratio < 0.8). The quantitative assay was rerun with diluted samples for values above the upper detection limit; 27 samples remained above the upper detection limit even after dilution. After the analysis, all samples were discarded.

##### Statistical analysis

The aim of the analyses was to assess the test characteristics of the IgG test assays based on DBS compared to serum samples and, if appropriate, to derive an adapted cutpoint and, in case of the quantitative S-antibody test, a potential correction formula for DBS results so that the seroprevalence and quantitative measures based on DBS are comparable to those based on serum samples. Results of the serum measurement were regarded as the gold standard for this analysis.

For the S-antibody test, a linear regression model was run for the log-transformed values in order to determine whether a correction formula was needed to predict quantitative serum values from DBS values. Values above the upper detection limit were set to the value of the upper detection limit. For sensitivity analysis, a model without the observations above the detection limit was run. Bland-Altman-Plots [4] were used to check for agreement visually. For this plot, the difference between the (log) DBS value and the (log) DBS value is plotted against the mean of the two (log) values.

For both the quantitative S-antibody and the semiquantitative N-antibody test, the categorised values were examined for agreement and equal marginal frequencies between serum and DBS classifications using McNemar's test [5].The categorisation used was 'positive' versus 'non-positive' (negative or indeterminate) using the manufacturer-supplied cutpoints. The categorisation for S-antibodies showed perfect agreement between serum and DBS results in our sample (see Supplemental Table S2), so further analysis of S-antibody misclassification was not necessary.

| Result of serum sample | Result of DBS | | Total |
|---|---|---|---|
| | Positive | Non-positive | |
| Positive | 241 (98.8 % | 0 (0 %) | 241 (98.8 %) |
| Non-Positive | 0 (0 %) | 3 (1.2 %) | 3 (1.2 %) |
| Total | 241 (98.8 %) | 3 (1.2 %) | 244 (100 %) |

**Supplemental Table S2:** Categorised IgG S-antibody measurement in serum vs. DBS using the manufacturer-supplied cutpoint, unweighted absolute and cell percentages

For the N-antibody results, an adapted cutpoint for DBS values was determined using the discordant proportion ratio [6], the ratio of the percentage of false positives to the percentage of false negatives. The null hypothesis of McNemar's test can be expressed as a discordant proportion ratio of 1. In our application this would indicate that the DBS result is not systematically biased towards false positives or false negatives, compared to the serum sample. For this purpose, cutpoints in the range of 0.80-1.50 were used to classify the DBS values. For each cutpoint, the proportion of misclassified DBS test results in comparison to serum results was determined and the ratio of false-positive to false-negative results was calculated. The cutpoint that led to the discordant proportion ratio closest to 1 was chosen as the adapted cutpoint.

In the analyses of the N-antibody results, weights were used to account for difference in seroprevalence (the proportion of positive N-antibody results) between the validation study and the main study. This procedure ensures that the results of the validation study are applicable to the main study, since it makes the marginal probabilities for positive and negative DBS test results identical to those observed in the main study. N-antibody positive observations were weighted with the ratio of the raw proportion of positive test results (5.2% in the main study to 17.6% in the validation study resulting in a weight <1) and negative ones with the respective proportions for negatives (94.8% in the main study, 82.4% in the validation study resulting in a weight >1). McNemar's test, however, was conducted on the unweighted data. As a further check of the adapted cutpoint, the weighted discordant proportion difference before and after cutpoint adaptation was tested against the null hypothesis of a difference equal to zero [7].

Confidence intervals for the proportion of misclassified DBS test results were calculated using the Wilson score method [7,8]. The logit method with survey procedures was used for the confidence intervals of weighted percentages.

#### Results

The linear regression models and Bland-Altman plots (see Supplemental Figure S2) indicated good agreement between serum and DBS values for the S-antibody test. For the S-antibody test, the explained variance ($R^2$) for the DBS values was 97.0%. The intercept (-0.078) had a standard error of 0.064 and was therefore not significantly different from zero. The slope parameter (1.012) had a standard error of 0.011 and was not significantly different from 1. The residual standard error was 0.205 on 242 degrees of freedom. Since neither the intercept nor the slope was significantly different from 0 or 1, respectively, no correction formula was derived. Due to a slightly skewed Bland-Altman plot for the full sample, the analysis was repeated for the S-antibody positive observations only (lower row of Supplemental Figure S2), showing good agreement. Excluding the observations above the upper detection limit did not change this result, either.

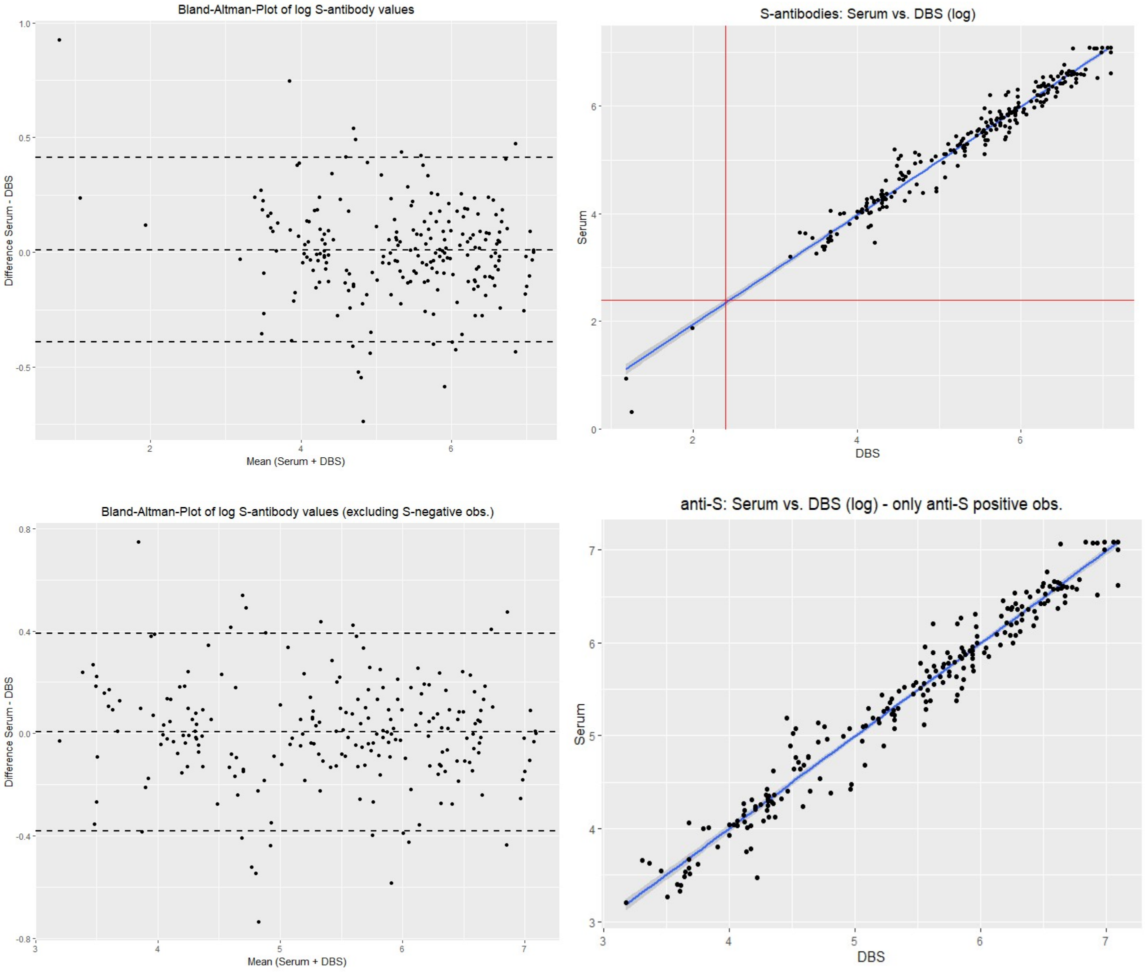

**Supplemental Figure S2: Left panel:** Bland-Altman plot of the difference between the quantitative serum log S1-IgG value and the corresponding DBS log value against the mean of the two log values. The dashed lines show the limits of agreement (± 1.96 standard deviations). Plots in the lower row exclude S-antibody negative observations. **Right panel**: Data points and regression line for the regression of quantitative serum log S1-IgG values on DBS log S1-IgG values. The red lines indicate the cutpoint between positive and non-positive categorisation.

For the categorised N-antibody test results, the proportion of misclassified DBS samples was 6.6% compared to the corresponding serum sample, applying the manufacturer-supplied cutpoint of 1.1 to the DBS samples (16 of 244 dried blood samples were misclassified, 95% CI 4.1-10.4%) (see Supplemental Table S3). There were both false negative and false positive categorisations whereby 12 (4.9%) positives in serum were false negatives (95% CI 2.8-8.4%) and 4 (1.6%) negatives in serum were false positives in DBS (95% CI 0.6-4.1%).

McNemar's test without continuity correction provided support against the null hypothesis of equal marginal frequencies (p=0.0455), suggesting a statistically significant difference between the serum and DBS classifications. The estimated difference of discordant proportions was 5.2% (p=0.0015), supporting this interpretation. Adapting the cutpoint for positivity for DBS values was therefore considered appropriate. The discordant proportion ratio closest to 1 was reached with a cutpoint of 0.95. When re-applying McNemar's test to the marginal frequencies after cutpoint adaptation, the null hypothesis could no longer be rejected on any meaningful significance level (p=0.4227). The estimated difference of discordant proportions then was at -0.5% (p=0.7478). The use of this cutpoint led to an unweighted overall misclassification of 5.7% (14 of 244 samples misclassified, 95% CI 3.4-9.4%) and false positive and false negative misclassifications occurring with similar weighted frequency of 2.8% and 2.4% (5.2%, 12.7 of 244 samples misclassified, 95% CI 3.0-9.0%) (see Supplemental Table S3).

| DBS - Cutpoint: 1.1 | | | |
|---|---|---|---|
| Serum / DBS | Positive | Non-Positive | Total |
| Positive | 39 (4.8%) | 4 (0.5%) | 43 (5.2%) |
| Non-Positive | 12 (5.7%) | 189 (89.1%) | 201 (94.8%) |
| Total | 51 (10.4%) | 193 (89.6%) | N=244 |
| **DBS - Cutpoint: 0.95** | | | |
| Serum / DBS | Positive | Non-Positive | Total |
| Positive | 46 (8.1%) | 9 (2.8%) | 55 (10.9%) |
| Non-Positive | 5 (2.4%) | 184 (86.7%) | 189 (89.1%) |
| Total | 51 (10.4%) | 193 (89.6%) | N = 244 |

**Supplemental Table S3:** Categorised IgG N-antibody measurement in serum vs. categorised IgG N-antibody measurement in dried blood spot using the manufacturer-supplied cutpoint (1.1) and adapted cutpoint (0.95), unweighted absolute numbers and weighted cell percentages

##### Implementation in the analysis of the seroprevalence study

For IgG S-antibodies, the values of the quantitative Euroimmun QuantiVac ELISA were not found to differ significantly between serum and dried blood samples. For IgG N-antibodies, however, an adapted cutpoint of 0.95 was obtained for classifying dried blood spot samples as N-antibody positive. This cutpoint was therefore used in the analysis of the RKI-SOEP-2 study presented here to classify the semiquantitative values of the Euroimmun IgG N-antibody test in dried blood spot samples.

### 3. Stratum-specific sensitivity estimates

Sensitivity was estimated internally from the study population. Here, we present the sensitivities estimated within categories of the stratification variables, for the stratified analyses of seroprevalence. For the logistic regression analysis (combined seroprevalence only), sensitivity was also estimated via a logistic model, with the same covariates as the main analysis (data not shown).

#### 3.1. Combined seroprevalence

The sensitivity for detecting combined seroprevalence was estimated by the proportion that was seropositive for anti-S or anti-N antibodies among study participants with a self-reported vaccination or a self-reported positive SARS-CoV-2 test at least 11 days pre-study (n = 9,260).

| | Anti-S or anti-N seropositive | | | | | |
|---|---|---|---|---|---|---|
| | Sensitivity | (95%-CI) | | N positive | *p*-value | N |
| **All (total 14-99 years)** | 95.8% | (95.1 - | 96.4) | 8915 | | 9260 |
| **All (total 18-99 years)** | 95.5% | (94.8 - | 96.2) | 8435 | | 8771 |
| **Women** | 96.3% | (95.4 - | 97.0) | 4820 | 0.1152 | 4980 |
| **Men** | 95.3% | (94.1 - | 96.2) | 4095 | | 4280 |

| | Anti-S or anti-N seropositive | | | | | |
|---|---|---|---|---|---|---|
| | Sensitivity | (95%-CI) | | N positive | *p*-value | N |
| **14-17 years** | 99.4% | (98.7 - | 99.8) | 480 | | 489 |
| **18-34 years** | 97.4% | (95.5 - | 98.5) | 1580 | | 1616 |
| **35-49 years** | 96.7% | (95.1 - | 97.8) | 2001 | | 2051 |
| **50-64 years** | 95.8% | (94.5 - | 96.9) | 2870 | | 2974 |
| **65-99 years** | 92.4% | (90.5 - | 93.8) | 1984 | <.0001 | 2130 |
| **Low deprivation** | 96.4% | (95.1 - | 97.4) | 2209 | | 2286 |
| **Medium deprivation** | 96.0% | (95.1 - | 96.8) | 5456 | | 5654 |
| **High deprivation** | 93.3% | (90.6 - | 95.2) | 1170 | 0.0107 | 1239 |
| **Northern region** | 94.5% | (91.9 - | 96.3) | 1637 | | 1696 |
| **Western region** | 95.5% | (94.2 - | 96.6) | 2848 | | 2960 |
| **Southern region** | 96.5% | (95.3 - | 97.4) | 2535 | | 2627 |
| **Eastern region** | 96.4% | (95.3 - | 97.3) | 1895 | 0.1564 | 1977 |

**Supplemental Table S4:** Stratum-specific sensitivity for detecting combined seroprevalence, estimated from the study population. Population-weighted and taking clustering within households into account in the estimation of confidence intervals.

#### 3.2. Infection-induced seroprevalence

The sensitivity for detecting infection-induced seroprevalence was estimated by the proportion that was seropositive for anti-N antibodies among participants with a self-reported positive SARS-CoV-2 test at least 11 days pre-study (n = 774).

| | Anti-N seropositive | | | | | |
|---|---|---|---|---|---|---|
| | Sensitivity | (95%-CI) | | N positive | *p*-value | N |
| **All (total 14-99 years)** | 47.4% | (41.6 - | 53.3) | 358 | | 774 |
| **All (total 18-99 years)** | 45.5% | (39.6 - | 51.5) | 322 | | 710 |
| **Women** | 65.2% | (45.4 - | 80.9) | 36 | | 64 |
| **Men** | 35.0% | (25.3 - | 46.1) | 61 | | 160 |
| **14-17 years** | 47.3% | (36.9 - | 57.9) | 103 | | 228 |
| **18-34 years** | 48.9% | (39.1 - | 58.8) | 113 | | 231 |
| **35-49 years** | 55.6% | (41.0 - | 69.2) | 45 | 0.0390 | 91 |
| **50-64 years** | 45.7% | (38.3 - | 53.4) | 191 | | 426 |
| **65-99 years** | 49.0% | (40.9 - | 57.3) | 167 | 0.5478 | 348 |
| **Low deprivation** | 51.3% | (38.3 - | 64.2) | 84 | | 177 |
| **Medium deprivation** | 44.3% | (37.5 - | 51.3) | 227 | | 495 |
| **High deprivation** | 59.6% | (45.8 - | 72.0) | 45 | 0.2190 | 95 |
| **Northern region** | 44.8% | (29.8 - | 60.8) | 40 | | 75 |
| **Western region** | 46.1% | (36.0 - | 56.5) | 88 | | 216 |
| **Southern region** | 48.3% | (37.2 - | 59.5) | 105 | | 229 |
| **Eastern region** | 49.1% | (38.7 - | 59.5) | 125 | 0.9658 | 254 |

**Supplemental Table S5:** Stratum-specific sensitivity for detecting infection-induced seroprevalence, estimated from the study population. Population-weighted and taking clustering within households into account in the estimation of confidence intervals.

### 4. Logistic regression with correction for test characteristics

For the analysis of combined seroprevalence with a logistic regression model, we used predictive value weighting to correct for test characteristics [1]. First, sensitivity for combined seroprevalence was estimated via a logistic model, with the same covariates as the main analysis (sex, age group, socio-economic deprivation, region and month of participation). This analysis was restricted to study participants with a self-reported vaccination or a self-reported infection at least 11 days pre-study (n = 9,260). The same model variables were used to estimate the uncorrected seroprevalence, and we applied the usual correction for sensitivity and specificity, using the model-based sensitivity estimates, to arrive at corrected seroprevalence estimates based on the model [2]. To obtain odds ratio estimates, we applied predictive value weighting. Predictive value weighting is based on the idea that each observed positive value is associated with a positive predictive value (PPV), i.e. the probability that the observed result is correct. (In the same way, each observed negative value comes with a negative predictive value.) The predictive values can be calculated from specificity, the model-based sensitivity and the corrected model-based seroprevalence according to standard formulae [3]. Each observed positive value is then true only with probability given by the positive predictive value (PPV), and is thus weighted with the PPV. With probability 1 – PPV, however, the true value is negative. Therefore, all positive observations are duplicated to form pseudo-observations. In the pseudo-observations, the observed "positive" is set to "negative" and the predictive value weight is given by 1 – PPV. Observed negative values are handled analogously, using the NPV. Finally, the predictive value weights are multiplied with the survey weights and used in the logistic regression analysis of the combined dataset of original and pseudo-observations with the assumed true antibody state as target variable. See the example in Supplemental Table S6.

| ID | Type of observation | Observed antibody result | True antibody state | Observed sero-prevalence* | Estimated sensitivity* | Corrected sero-prevalence* | Predictive value weight* | Survey weight | Weight for logistic model |
|---|---|---|---|---|---|---|---|---|---|
| **1** | Original | Positive | Positive | 0.900 | 0.967 | 0.931 | PPV = 0.9995 | 0.81 | 0.8096 |
| **1** | Pseudo | Positive | Negative | 0.900 | 0.967 | 0.931 | 1 – PPV = 0.0005 | 0.81 | 0.0004 |
| **2** | Original | Negative | Negative | 0.900 | 0.967 | 0.931 | NPV = 0.690 | 1.33 | 0.9177 |
| **2** | Pseudo | Negative | Positive | 0.900 | 0.967 | 0.931 | 1 – NPV = 0.310 | 1.33 | 0.4123 |

**Supplemental Table S6:** Calculation of predictive value weights for two hypothetical observations from the same group (e.g. female, 35-49 yrs, district with medium deprivation, western region, participation in November).
PPV = positive predictive value, NPV = negative predictive value, specificity 0.994.
*Values are identical for all observations from the same group.

95% confidence intervals for the odds ratios and the adjusted prevalences derived from the model were estimated via a bootstrap procedure with 2000 replications, bootstrapping the whole process starting from the model-based estimation of sensitivity, so that the uncertainty stemming from estimating the sensitivity internally is included in the confidence intervals as well as the uncertainty from estimating the predictive values. The bootstrap replicates were generated through random sampling with replacement from the original data, using the households as sampling units. Confidence intervals were determined by the percentile method (P2.5 and P97.5).

#### 5. Estimation of the underreporting factor

Underreporting of infections in COVID-19 case notifications was estimated by linking the study data to mandatory notification data [1]. To this end, we calculated the cumulative proportion of notified COVID-19 cases (excluding deceased cases) from the start of the pandemic until each day of the field period within strata defined by age group, sex and district. This prevalence of notified cases over the first two years of the pandemic was matched to participants by DBS sampling date (minus 14 days difference to symptom onset), age group, sex and district. This prevalence was then averaged over all participants in the study (or over all participants in a stratum, for a stratified analysis) to yield an estimate of the cumulative proportion of non-fatal cases in COVID-19 case notification data. The underreporting factor was calculated as the ratio of infection-induced seroprevalence in the study to the cumulative proportion of notified cases. Hereby, infection-induced seroprevalence was defined as seropositivity for anti-N antibodies, with correction for test characteristics. Confidence intervals for the underreporting factor were obtained by dividing the CI limits for the corrected seroprevalence by the cumulative proportion of notified cases. $p$-values for comparing categories of a stratification variable were based on the difference of the log ratios [2].

### 6. Comparison with national surveillance data

In this section, the study population is compared with national surveillance data, regarding vaccination/basic immunization and regarding known infections.

#### 6.1. Comparison with national vaccination monitoring

Supplemental Figure S3 compares the population-weighted prevalence of basic immunization in the study with the coverage of basic immunization derived from national surveillance data (Digital Vaccination Coverage Monitoring, DIM [1]). Data in the vaccination monitoring derive from vaccination centres and doctors, capturing the aggregated number of daily vaccinations, grouped by type of vaccination (first vaccination vs. basic immunization vs. booster), federal state (place of vaccination) and age group. Vaccination coverage is only available for pre-defined age groups, thus the comparison is only possible for two adult age groups (18 to 59 years, 60 years and older).

The DIM vaccination coverage shown in the figure was derived by assigning the vaccination coverage to each participant in the data set according to his or her participation date (counting all vaccinations reported until the participation date), federal state (place of living) and age group. A weighted average of the assigned vaccination coverage over each age group then represents the DIM vaccination coverage for this group, considering the different dates of study participation and the composition of the survey sample with respect to federal states (which exhibit variation in vaccination coverage between them).

In contrast to the main part of the paper, basic immunization here only considers vaccinations and known infections, not antibodies. Thus, comparability of the study data with the vaccination monitoring is achieved. As only infections before a vaccination can be considered in the DIM vaccination monitoring (because data in the reporting system are updated when vaccination occurs), the variable used here also only considers infections prior to a vaccination. Moreover, our definition of basic immunization with the JCOVDEN vaccine (previously COVID-19 Vaccine Janssen) was adapted here to the definition adopted by the DIM at that time, therefore, a single dose of JCOVDEN was considered sufficient in this case to be categorized as having a basic immunization.

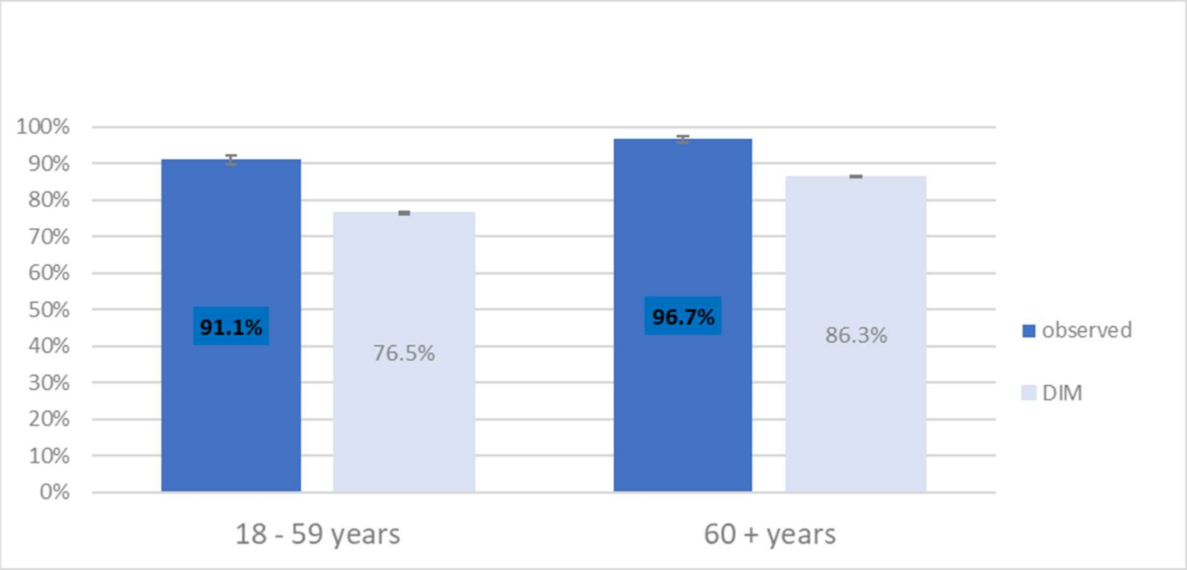

**Supplemental Figure S3.** Comparison of prevalence of basic immunization observed in the RKI-SOEP-2 study with national vaccination monitoring (DIM). Definition of basic immunization in the study data adapted to information available in the vaccination monitoring

Among study participants, the observed weighted prevalence of basic immunization in the age group 18 to 59 years was 91.1% (95% CI, 89.8%–92.3%), and 96.7% (95.6%–97.5%) in the age group 60 years and older. The matched basic immunization rates according to the vaccination monitoring, however, are 76.5% (95% CI, 76.2%-76.7%) for 18 to 59 years, and 86.3% (95% CI, 86.2%-86.5%) for 60 years and older. Observed rates in our study were thus 10 to 15 percentage points higher. This points to the presence of some selection bias in our study, with health-conscious individuals and persons that are in agreement with pandemic mitigation measures and vaccination being more likely to participate, leading to an overrepresentation of vaccinated individuals in the sample. But there are other factors contributing to this difference. The vaccination monitoring may not capture all vaccinations, and therefore underestimate the true vaccination coverage in the population [2]. Furthermore, the comparison is hampered by the fact that there are only very broad age groups available in the vaccination monitoring, with possible strong variation in vaccination coverage by age within these age groups, as vaccination priorisation heavily depended on age. Moreover, vaccinations are partly reported with a considerable time lag [2], which we did not account for in our analysis. Therefore, we consider the observed difference to basic immunization rates observed in the DIM vaccination monitoring as an upper boundary for the extent of selection bias in our study.

#### 6.2. Comparison with mandatory COVID-19 case notification

The self-reported PCR-confirmed infections in the study correspond to COVID-19 cases reported in the mandatory notification system in Germany [3]. Therefore, they can directly be compared to notification data in order to assess possible selection bias in the study. Supplemental Figure S4 shows the proportion with known, self-reported infection in the study (observed proportion) compared to the cumulative proportion of non-fatal cases in the German population according to notification data (expected proportion). To calculate the expected proportion in the population, the infection rate based on notified cases (number of non-fatal cases, divided by population size minus the number of fatal cases) was matched to the individual participants by participation date (minus 4 days difference to symptom onset), age group and district (similarly to the estimation of underreporting, see Supplement 4, but with a different time lag). This adjusts for changes in the cumulative proportion over time and differences between districts and age groups. The expected proportion for each group was obtained by calculating groupwise weighted averages of the individually matched infection rates.

The proportion with known infection observed in the study was 8.3% (95% CI 5.8%-11.9%) for 14 to 17 years, 8.0% (95% CI 6.4%-9.9%) for 18 to 34 years, 11.6% (95% CI 9.5%-14.0%) for 35 to 49 years, 6.5% (95% CI 5.3%-7.9%) for 50 - 64 years and 4.0% (95% CI 3.1%-5.1%) for 65 years and older. The expected proportion based on notification data was 11.1% (95% CI 10.5%-11.7%) for 14 to 17 years, 10.4% (95% CI 10.1%-10.6%) for 18 to 34 years, 9.4% (95% CI 9.1%-9.7%) for 35 to 49 years, 6.5% (95% CI 6.4%-6.7%) for 50 - 64 years and 3.9% (95% CI 3.8%-4.0%) for 65 years and older. In total, the observed proportion was 7.4% (95% CI 6.6%-8.3%), only slightly lower than the expected proportion of 7.7% (95% CI 7.6%-7.9%).

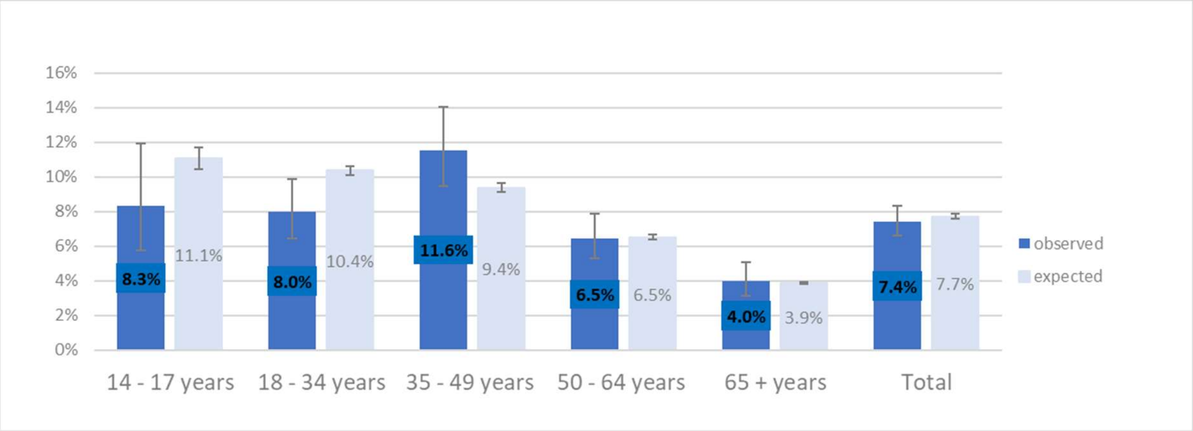

**Supplemental Figure S4.** Comparison of prevalence of known SARS-CoV-2 infections estimated from RKI-SOEP-2 study (observed) with prevalence of notified cases from surveillance data (expected).

In the total and in the higher age groups (50 years and older), there was good agreement between the observed and expected proportion of known infections. In the youngest two age groups (up to 34 years), however, participants were less likely to report a known infection than the general population, while in the age group 35 to 49 years the proportion observed in the study was higher than expected in the population. A possible explanation for the lower observed infection rate in the youngest age groups might be that teenagers and young adults are generally healthy and not that much inclined to take part in a health study, whereas the subgroup of young people with health problems might be more interested in participating in the study, while at the same time having protected themselves more against infection than their peers since they were more vulnerable. Older age groups, in contrast, had higher response rates in our study [4], and at the same time a higher proportion of vulnerable individuals, so there is less potential for selection bias.

This analysis suggests that, as a whole, our sample is similar to the general population with regard to known infections, however, there is some selective participation in the three younger age groups.

#### 7. Results stratified by federal state

Here, we present analyses of combined seroprevalence, SARS-CoV-2 infection status and basic immunization stratified by federal state. Results for infection-induced seroprevalence and the underreporting factor are not shown due to small sample size. Also, we did not perform the logistic regression analysis for combined seroprevalence with correction for sensitivity and specificity, because it produced large confidence intervals in the bootstrap analysis.

For this analysis, the sampling weights were additionally adjusted for the age distribution within each federal state. After controlling for multiple comparisons (due to the high number of 16 states being compared), we observed a lower combined seroprevalence and basic immunization rate in Saxony and Brandenburg (combined seroprevalence: 89.1% and 87.4% vs. 95.2% overall, with correction for specificity and stratum-specific sensitivity; basic immunization: 81.4% and 80.5% vs. 90.7% overall) and a higher infection rate in Saxony (18.5% vs. 9.8% overall). The difference in the prevalence of basic immunization was somewhat smaller in the model-adjusted analysis (basic immunization: 82.4% and 80.9% vs. 90.7% overall).

| | Total | N+* | S+* | Combined Seroprevalence (N+ or S+), uncorrected | | | | Combined Seroprevalence (N+ or S+), corrected for specificity and stratum-specific sensitivity |
|---|---|---|---|---|---|---|---|---|
| | N (column %) | %** | %** | N positive | %** | *p*-value (each state vs. the others) **** | *p*-value (each state vs. the others), corrected for multiple testing**** | Prevalence***, % (95% CI) |
| All (total 14-99 years) | 10687 | 6.0 | 90.4 | 9795 | 91.1 (90.1-91.9) | | | 95.2 (94.0-96.3) |
| **Federal State** | | | | | *p = 0.0008* | | | |
| Schleswig-Holstein | 423 (3.8) | 4.1 | 92.7 | 395 | 92.8 (88.3-95.7) | 0.38 | 1.00 | 95.2 (90.9-99.5) |
| Hamburg | 238 (2.2) | 7.7 | 90.7 | 223 | 90.7 (81.6-95.5) | 0.90 | 1.00 | 93.3 (85.7-101.0) |
| Lower Saxony | 1005 (9.5) | 3.7 | 90.4 | 936 | 90.5 (86.7-93.3) | 0.72 | 1.00 | 96.9 (92.0-101.7) |
| Bremen***** | 67 (0.8) | | | | | | | |
| North Rhine-Westphalia | 2143 (21.2) | 4.6 | 93.1 | 2016 | 93.2 (91.2-94.8) | 0.019 | 0.26 | 96.9 (94.6-99.2) |
| Hesse | 733 (7.8) | 6.9 | 91.3 | 676 | 91.7 (87.9-94.3) | 0.71 | 1.00 | 96.9 (92.7-101.1) |
| Rhineland-Palatinate | 283 (4.1) | 4.6 | 89.7 | 352 | 89.7 (83.3-93.9) | 0.58 | 1.00 | 95.6 (88.5-102.6) |
| Baden-Württemberg | 1322 (13.8) | 5.6 | 91.8 | 1217 | 91.9 (88.9-94.1) | 0.50 | 1.00 | 95.8 (92.5-99.1) |
| Bavaria | 1679 (15.7) | 6.4 | 91.8 | 1546 | 92.8 (90.8-94.4) | 0.065 | 0.77 | 96.0 (93.8-98.2) |
| Saarland***** | 109 (1.5) | | | | | | | |
| Berlin | 514 (5.0) | 5.0 | 91.7 | 477 | 91.6 (86.5-94.9) | 0.79 | 1.00 | 93.1 (88.8-97.4) |
| Brandenburg | 406 (2.5) | 9.0 | 83.3 | 357 | 83.8 (77.5-88.6) | 0.0008 | 0.012 | 87.4 (80.9-93.9) |
| Mecklenburg-Pomerania | 227 (1.6) | 3.2 | 85.5 | 196 | 85.5 (76.8-91.3) | 0.057 | 0.75 | 89.5 (81.3-97.7) |
| Saxony | 726 (5.3) | 12.7 | 83.0 | 621 | 83.3 (78.0-87.5) | <.0001 | 0.0003 | 89.1 (83.3-94.8) |
| Saxony-Anhalt | 334 (2.6) | 8.7 | 87.4 | 298 | 88.8 (82.9-92.8) | 0.31 | 1.00 | 92.5 (86.6-98.3) |
| Thuringia | 378 (2.7) | 7.1 | 88.0 | 322 | 88.1 (82.7-92.0) | 0.16 | 1.00 | 92.0 (86.2-97.8) |

**Supplemental Table S7:** Characteristics and combined IgG seroprevalence in community-dwelling persons (≥14 years) in Germany (10,687 RKI-SOEP study participants with valid dried blood spot specimens, sampled predominantly between November 2021-January 2022). All percentages are population-weighted (weights additionally adjusted for the age distribution within federal states).
*N+, anti-N seropositive. S+, anti-S seropositive. Number of available cases for anti-N-antibodies result: 10,342; anti-S-antibodies result: 10,687.
**Percentage without correction for sensitivity and specificity.
***Prevalence for (N+ and/or S+), corrected for sensitivity (including antibody waning, internally estimated, stratum-specific values, see Supplement 3) and anti-S specificity = 0.994.
****$p$-value for comparing each state with the other states (based on the unadjusted prevalence estimate), corrected for multiple testing (16 states) using the Bonferroni-Holm method (ref. Holm 1979 [1].

| | Total | self-reported PCR+* | N+* | unvaccinated & S+* | SARS-CoV-2 Infection | | | | | |
|---|---|---|---|---|---|---|---|---|---|---|
| | N (column %) | % | % | % | N positive | Prevalence, % (95% CI) | *p*-value (each state vs. the others) **** | *p*-value (each state vs. the others), corrected for multiple testing**** | OR and *p*-value, adjusted** (95% CI) | Model-adjusted** prevalence, % (95% CI) |
| All (total 14-99 years) | 11154 | 7.3 | 5.9 | 1.3 | 1193 | 9.8 (8.9-10.7) | | | | 9.7 (8.8-10.6) |
| **Federal State** | | | | | | | | | *p < 0.0001* | |
| Schleswig-Holstein | 436 (3.7) | 5.1 | 4.1 | 0.1 | 29 | 6.7 (3.4-12.6) | 0.237 | 1.00 | 0.88 (0.41-1.89) | 7.3 (3.8-13.8) |
| Hamburg | 248 (2.3) | 4.8 | 7.7 | 0.0 | 18 | 9.4 (4.9-17.4) | 0.903 | 1.00 | 1.15 (0.52-2.55) | 9.3 (4.8-17.3) |
| Lower Saxony | 1042 (9.4) | 4.9 | 3.7 | 0.9 | 71 | 6.7 (4.4-10.1) | 0.056 | 0.73 | 0.82 (0.49-1.35) | 6.8 (4.5-10.2) |
| Bremen*** | 71 (0.8) | | | | | | | | | |
| North Rhine-Westphalia | 2245 (21.4) | 6.6 | 4.6 | 0.7 | 205 | 8.3 (6.7-10.4) | 0.11 | 1.00 | Ref. | 8.2 (6.5-10-3) |
| Hesse | 754 (7.7) | 8.5 | 6.9 | 0.8 | 77 | 11.1 (7.7-15.6) | 0.47 | 1.00 | 1.37 (0.86-2.21) | 10.9 (7.6-15.4) |
| Rhineland-Palatinate | 410 (4.3) | 5.8 | 4.6 | 0.7 | 39 | 7.0 (3.9-12.1) | 0.23 | 1.00 | 0.80 (0.41-1.57) | 6.7 (3.8-11.8) |
| Baden-Württemberg | 1385 (13.7) | 6.7 | 5.6 | 2.2 | 146 | 9.8 (7.4-12.9) | 0.97 | 1.00 | 1.19 (0.77-1.83) | 9.6 (7.1-12.7) |
| Bavaria | 1761 (15.8) | 7.6 | 6.4 | 1.4 | 196 | 9.7 (7.6-12.3) | 0.97 | 1.00 | 1.18 (0.79-1.77) | 9.6 (7.4-12.3) |
| Saarland*** | 111 (1.5) | | | | | | | | | |
| Berlin | 537 (5.0) | 10.2 | 5.0 | 1.6 | 71 | 12.3 (8.7-17.1) | 0.17 | 1.00 | 1.53 (0.95-2.46) | 11.9 (8.3-16.9) |
| Brandenburg | 423 (2.5) | 12.1 | 9.0 | 2.8 | 61 | 15.5 (10.2-22.7) | 0.026 | 0.37 | 1.97 (1.20-3.24) | 14.8 (10.2-20.9) |
| Mecklenburg-Pomerania | 233 (1.6) | 4.8 | 3.2 | 0.2 | 11 | 5.8 (2.8-11.8) | 0.15 | 1.00 | 0.76 (0.32-1.80) | 6.4 (3.0-13.5) |
| Saxony | 756 (5.3) | 14.2 | 12.7 | 4.5 | 148 | 18.5 (14.3-23.5) | <.0001 | <.0001 | 2.81 (1.89-4.17) | 19.6 (15.2-24.9) |
| Saxony-Anhalt | 354 (2.6) | 6.3 | 8.7 | 0.2 | 46 | 12.0 (7.4-18.9) | 0.39 | 1.00 | 1.57 (0.82-3.02) | 12.2 (7.1-20.2) |
| Thuringia | 388 (2.7) | 7.1 | 7.1 | 2.3 | 61 | 11.0 (7.5-15.8) | 0.53 | 1.00 | 1.40 (0.85-2.32) | 11.1 (7.5-16.1) |

**Supplemental Table S8:** Characteristics and SARS-CoV-2 infection status in community-dwelling persons (≥14 years) in Germany (11,154 RKI-SOEP-2 study participants, predominantly November 2021-January 2022). All percentages are population-weighted (weights additionally adjusted for the age distribution within federal states).

*N+, anti-N seropositive. S+, anti-S seropositive. Number of available cases for self-reported PCR test: 10,889; anti-N-antibodies result: 10,342; self-reported vaccination status & anti-S-antibodies result: 10,501.

**Model variables: sex, age group, district-level socio-economic deprivation, federal state, month of study participation (categorical variable). *p*-value for a Wald test of each variable within a survey logistic regression model.

***Results not shown due to small sample size.

****$p$-value for comparing each state with the other states (based on the unadjusted prevalence estimate), corrected for multiple testing (16 states) using the Bonferroni-Holm method [1].

| | Total | at least two doses of vaccine* | hybrid immunity* | Basic Immunization | | | | | |
|---|---|---|---|---|---|---|---|---|---|
| | N (column %) | % (95% CI) | % (95% CI) | N positive | Prevalence, % (95% CI) | *p*-value (each state vs. the others) **** | *p*-value (each state vs. the others), corrected for multiple testing **** | OR and *p*-value, adjusted** (95% CI) | Model-adjusted** prevalence, % (95% CI) |
| All (total 14-99 years) | 10932 | 88.8 (87.8-89.8) | 7.5 (6.8-8.4) | 9975 | 90.7 (89.7-91.6) | | | | 90.7 (89.7-91.6) |
| **Federal State** | | | | | | | | *p* < 0.0001 | |
| Schleswig-Holstein | 431 (3.6) | 90.3 (84.5-94.1) | 5.0 (2.4-10.1) | 400 | 90.8 (85.0-94.5) | 0.97 | 1.00 | 0.72 (0.38-1.36) | 91.4 (86.0-94.8) |
| Hamburg | 241 (2.2) | 88.1 (76.4-94.4) | 9.5 (4.9-17.7) | 223 | 88.9 (77.1-95.0) | 0.65 | 1.00 | 0.39 (0.15-1.03) | 85.6 (71.7-93.3) |
| Lower Saxony | 1020 (9.5) | 91.0 (87.2-93.8) | 4.6 (3.1-6.7) | 961 | 92.1 (88.4-94.7) | 0.38 | 1.00 | 0.87 (0.51-1.47) | 92.7 (89.2-95.1) |
| Bremen*** | 65 (0.8) | | | | | | | | |
| North Rhine-Westphalia | 2197 (21.4) | 91.1 (88.7-93.0) | 7.1 (5.6-9.0) | 2076 | 93.2 (91.0-94.8) | 0.012 | 0.16 | Ref. | 93.6 (91.5-95.1) |
| Hesse | 733 (7.6) | 90.5 (86.8-93.3) | 9.3 (6.3-13.4) | 683 | 93.0 (89.7-95.4) | 0.13 | 1.00 | 0.88 (0.51-1.52) | 92.8 (89.4-95.2) |
| Rhineland-Palatinate | 407 (4.3) | 89.6 (82.9-93.8) | 6.3 (3.4-11.4) | 372 | 91.5 (85.6-95.1) | 0.73 | 1.00 | 0.75 (0.38-1.48) | 91.7 (85.9-95.2) |
| Baden-Württemberg | 1359 (13.7) | 89.4 (86.1-92.0) | 6.7 (4.8-9.4) | 1234 | 90.4 (87.1-92.9) | 0.84 | 1.00 | 0.57 (0.34-0.95) | 89.4 (85.4-92.5) |
| Bavaria | 1731 (15.8) | 90.2 (87.7-92.2) | 6.9 (5.2-9.1) | 1587 | 91.4 (89.0-93.3) | 0.49 | 1.00 | 0.61 (0.39-0.95) | 90.0 (87.1-92.4) |
| Saarland*** | 108 (1.4) | | | | | | | | |
| Berlin | 527 (5.0) | 88.1 (82.2-92.2) | 10.7 (7.5-15.1) | 497 | 92.0 (86.6-95.3) | 0.57 | 1.00 | 0.91 (0.47-1.78) | 93.0 (88.2-96.0) |
| Brandenburg | 411 (2.5) | 78.5 (70.9-84.5) | 9.5 (6.1-14.6) | 352 | 80.5 (72.8-86.4) | <.0001 | 0.0008 | 0.28 (0.16-0.48) | 80.9 (73.4-86.7) |
| Mecklenburg-Pomerania | 230 (1.6) | 88.8 (81.5-93.5) | 5.7 (2.7-11.8) | 206 | 90.3 (83.1-94.6) | 0.88 | 1.00 | 0.70 (0.33-1.49) | 91.1 (83.7-95.3) |
| Saxony | 745 (5.3) | 77.0 (71.2-82.0) | 12.4 (9.1-16.6) | 608 | 81.4 (75.9-85.9) | <.0001 | <.0001 | 0.31 (0.19-0.49) | 82.4 (77.3-86.6) |
| Saxony-Anhalt | 344 (2.6) | 85.0 (78.0-90.1) | 10.6 (6.2-17.5) | 302 | 87.0 (80.2-91.7) | 0.14 | 1.00 | 0.49 (0.25-0.95) | 88.0 (80.2-93.0) |
| Thuringia | 383 (2.7) | 87.1 (81.7-91.1) | 8.2 (5.2-12.8) | 315 | 88.1 (82.8-91.9) | 0.20 | 1.00 | 0.48 (0.27-0.83) | 87.7 (81.9-91.8) |

**Supplemental Table S9:** Characteristics and basic immunization status in community-dwelling persons (≥14 years) in Germany (10,932 RKI-SOEP-2 study participants, predominantly November 2021-January 2022). All percentages are population-weighted (weights additionally adjusted for the age distribution within federal states).
*Number of available cases for at least two doses of vaccine: 10,932; hybrid immunity: 10,925.
**Model variables: sex, age group, district-level socio-economic deprivation, federal state, month of study participation (categorical variable). *p*-value for a Wald test of each variable within a survey logistic regression model.
***Results not shown due to small sample size.
****p-value for comparing each state with the other states (based on the unadjusted prevalence estimate), corrected for multiple testing (16 states) using the Bonferroni-Holm method [1].
